# Brain development mutations in the β-tubulin TUBB result in defective ciliogenesis

**DOI:** 10.1101/2023.05.23.23290232

**Authors:** Antonio Mollica, Safia Omer, Sonia L. Evagelou, Serhiy Naumenko, Lu Yi Li, Aideen Teeling, Kyle Lindsay, Steven Erwood, Robert M. Vernon, Julie D. Forman-Kay, Manohar Shroff, Rene E. Harrison, Ronald D. Cohn, Evgueni A. Ivakine

## Abstract

Tubulinopathies and neurodevelopmental ciliopathies are two groups of genetic disorders characterized by abnormal brain development resulting in structural brain malformations. Tubulinopathies are caused by dominant missense mutations in genes encoding for tubulins, the building blocks of microtubules. Neurodevelopmental ciliopathies are mostly recessive disorders caused by defects in the function of the primary cilium, a sensory organelle that modulates signaling pathways important for brain development. Though more than 40 genes have been associated with neurodevelopmental ciliopathies, many patients still do not have an identified genetic etiology. Herein, we present a novel *de novo* heterozygous missense variant in Tubulin Beta Class I (*TUBB*) identified through whole-genome sequencing analysis in a patient with both ciliopathy and tubulinopathy brain features. While microtubules are fundamental to primary cilia formation and function, no association between mutations in tubulin genes and neurodevelopmental ciliopathies has been found to date. Using patient-derived cells and gene-edited isogenic cell lines, we show that the identified variant impairs the early stages of cilia formation by altering microtubule dynamics and structure. Furthermore, we demonstrate that the disease mechanism is not haploinsufficiency and that other patient mutations in *TUBB* affect cilia formation *in vitro*, putting forward defective ciliogenesis as a contributing pathogenic factor in a subset of tubulinopathy patients.

## INTRODUCTION

Genetic neurodevelopmental disorders are a diverse group of diseases characterized by abnormal development of the brain during embryogenesis, fetal development, or the early postnatal period^1,2^. These disorders are traditionally grouped into genetically heterogeneous classes of diseases that are either based on similarities in clinical and diagnostic features or based on the function of the relevant genes. Two groups of genetic neurodevelopmental disorders are the tubulinopathies and neurodevelopmental ciliopathies.

Tubulinopathies are a group of disorders characterized by cognitive and motor impairment, caused by dominant mutations in genes encoding for tubulins^3–6^. Tubulins are the building blocks of microtubules (MTs), one of the major components of the eukaryotic cytoskeleton. MTs are polymers of α- and β-tubulin heterodimers arranged head-to-tail to form protofilaments which interact longitudinally to create the hollow cylindrical structure of MTs^7^. In humans there are nine α- and nine β-tubulin isotypes, each encoded by a separate gene. Despite their high homology, there is increasing experimental and clinical evidence that the different tubulin isotypes are functionally distinct and not fully interchangeable^3,8,9^. The nine α- and nine β-tubulins are differentially expressed in different tissues at different times, generating a diverse repertoire of αβ dimers that confer distinct properties to the resulting MTs^10^. MTs are known to play a role in key neurodevelopmental processes such as neurogenesis, neuronal migration, axon formation and synaptogenesis^3,5^. Due to the importance of MTs for brain development, life-compatible tubulin mutations found in patients typically have subtle effects on MT properties and functions and do not severely compromise the overall functionality of MTs^4^. To date, all known pathogenic variants in tubulin genes are heterozygous missense mutations arising *de novo* in the affected individuals. Of the over twenty tubulin genes encoded by the human genome, six have been associated to tubulinopathies: one encoding α-tubulin (*TUBA1A*), four encoding β-tubulins (*TUBB*, *TUBB2A*, *TUBB2B*, *TUBB3*), and the γ-tubulin gene *TUBG1*^4^. Tubulinopathies are characterized by an array of complex brain malformations visible on brain imaging, typically affecting the cortex and other brain structures such as basal ganglia, cerebellum, and brainstem^4,5^. Tubulinopathies caused by mutations in different tubulin genes generally present with characteristic features, even though there is extensive phenotypic overlap^3–5^. Importantly, the molecular mechanisms underlying each structural malformation have not been fully elucidated.

Neurodevelopmental ciliopathies comprise Joubert syndrome (JS) and Meckel syndrome (MKS)^11,12^. Traditionally considered as two separate disorders due to their distinct phenotypes, JS and MKS significantly overlap in their causative genes and have recently been proposed to represent the mild (JS) and severe (MKS) ends of the same pathological spectrum^13^. JS and MKS belong to a group of diseases called ciliopathies because the pathogenic mutations affect the structure or the function of the primary cilium, a single-copy organelle present in most mammalian cells including neurons and neuronal progenitors^13,14^. Emerging as a hair-like protrusion of the plasma membrane, the primary cilium is a sensory organelle enriched in receptors and signaling factors^15^. Despite being continuous with the plasma membrane, it has a distinct protein and lipid composition that is maintained by a diffusion barrier at the ciliary base and by active transport mechanisms in and out of the cilium^16^. More than just an accumulation site for signaling molecules, the primary cilium plays an active role in modulating signal transduction by concentrating or excluding specific factors in a regulated fashion. In particular, the primary cilium regulates numerous signalling pathways that are important for embryogenesis, brain development and adult tissue homeostasis such as Hedgehog and Wnt^15,17^. Consistent with the broad role of the primary cilium, most ciliopathies can present in some patients with symptoms in various organ systems including the brain^11,12^. However, in the case of JS and MKS, brain malformations are defining and diagnostic features. JS originates from defective embryogenesis of midbrain and hindbrain structures, and affects approximately 1 in 100000 live births^18^. It is almost exclusively recessive, but an autosomal dominant mild form of JS has recently been described^13,19,20^. The distinctive diagnostic feature of JS is a cerebellar and brainstem malformation visible on brain imaging, referred to as the molar tooth sign (MTS). The MTS consists of cerebellar vermis hypoplasia or dysplasia, thick horizontal and elongated superior cerebellar peduncles, and a deep interpeduncular fossa^13,18^. At the cellular level, this characteristic set of brain malformations has been linked to defects in neurogenesis, neuronal migration, and axon guidance^13,17^. Clinically, JS typically presents with hypotonia evolving to ataxia, developmental delay, oculomotor apraxia and breathing abnormalities. In addition to the hallmark neurological symptoms, patients with JS may suffer from an array of symptoms in other organ systems such as kidney dysfunction, liver fibrosis and retinal defects^13,18^. MKS is a recessive condition that affects approximately 1 in 135000 live births^21^. It is the most severe ciliopathy, causing pre- or perinatal lethality due to developmental defects in multiple organs. MKS is associated to a highly variable phenotype, but the most common and distinctive features are occipital encephalocele, enlarged cystic kidneys and liver fibrosis. Neurodevelopmental defects such as occipital encephalocele or other less common brain malformations are obligate features for MKS, which may also present with additional defects such as postaxial polydactyly, coloboma, cleft lip and palate, congenital heart defects, ambiguous genitalia, and skeletal abnormalities^13,21^. Despite having greater than 40 JS- and 15 MKS-associated genes identified thus far, the exact genetic basis of disease for many patients suffering from neurodevelopmental ciliopathies remains undetermined^13,22^.

In cell biology, the link between MTs and primary cilia is well established. Nine circularly arranged bundles of two MTs (doublets) make up the axoneme of primary cilia, a specialized structure that supports the cilium and allows for selective transportation of signaling factors in and out of the cilium^16^. Moreover, MTs are essential for the biogenesis of the primary cilium, which is assembled via the ciliogenesis pathway in the G1 phase of the cell cycle or in G0 following cell cycle exit^23^. In particular, neurons and neuronal progenitors rely on the intracellular pathway of ciliogenesis^24–27^. In intracellular ciliogenesis, endosomal and Golgi pre-ciliary vesicles (PCVs) are recruited to the mother centriole via both dynein- and myosin-mediated transport on MTs and actin filaments, respectively^28–30^. PCVs then fuse to form the ciliary membrane, initiating the growth of the ciliary axoneme while the centrosome/forming cilium complex moves apically towards the plasma membrane^30–32^. This movement is primarily mediated by pushing forces from dense MT bundles assembling against the cell cortex^32^. Despite this tight connection, no association between mutations in tubulin genes and primary ciliopathies has been found to date, and tubulinopathies and neurodevelopmental ciliopathies are considered as two independent groups of disorders. Additionally, the molecular mechanisms underlying tubulinopathies are poorly understood and many neurodevelopmental ciliopathies patients lack a genetic etiology.

In this work we investigate the potential role of primary cilia defects in the pathogenesis of tubulinopathies. Starting from a patient manifesting clinical features of both JS and tubulinopathy, we show that a subset of dominant brain development mutations in the β-tubulin TUBB affect primary cilia formation *in vitro,* revealing a novel disease mechanism for TUBB-related tubulinopathies and a novel ciliopathy-associated gene.

## RESULTS

### Patient with a neurodevelopmental disorder shows atypical, ciliopathy- and tubulinopathy-like brain MRI features

A male patient, the sole in his pedigree, presented from an early age with neurodevelopmental delay and various neurological symptoms including mild intellectual disability, ataxia, and oculomotor apraxia. The patient displayed only neurological symptoms without involvement of other organs or organ systems.

Magnetic resonance imaging (MRI) of the patient’s brain **(Figure 1a)** revealed features compatible with JS, i.e., superior vermis hypoplasia, mildly long, thick, and horizontal superior cerebellar peduncles, and elevated roof of the 4^th^ ventricle, which show up as the classic MTS on axial images **(Figure 1a-ii)**. In addition, the MRI showed other abnormalities such as dysplastic somewhat asymmetric appearance of the superior cerebellar hemispheres **(Figure 1a-iii)** and asymmetric, somewhat bulbous and abnormal caudate nucleus on the right **(Figure 1a-iv and 1a-v)**, which are soft features of tubulinopathy^5^. Other findings were also present, such as hypoplastic olfactory bulbs **(Figure 1a-vi)** and incomplete rotation of the left hippocampus **(Figure 1a-vii)**.

**Figure 1:**
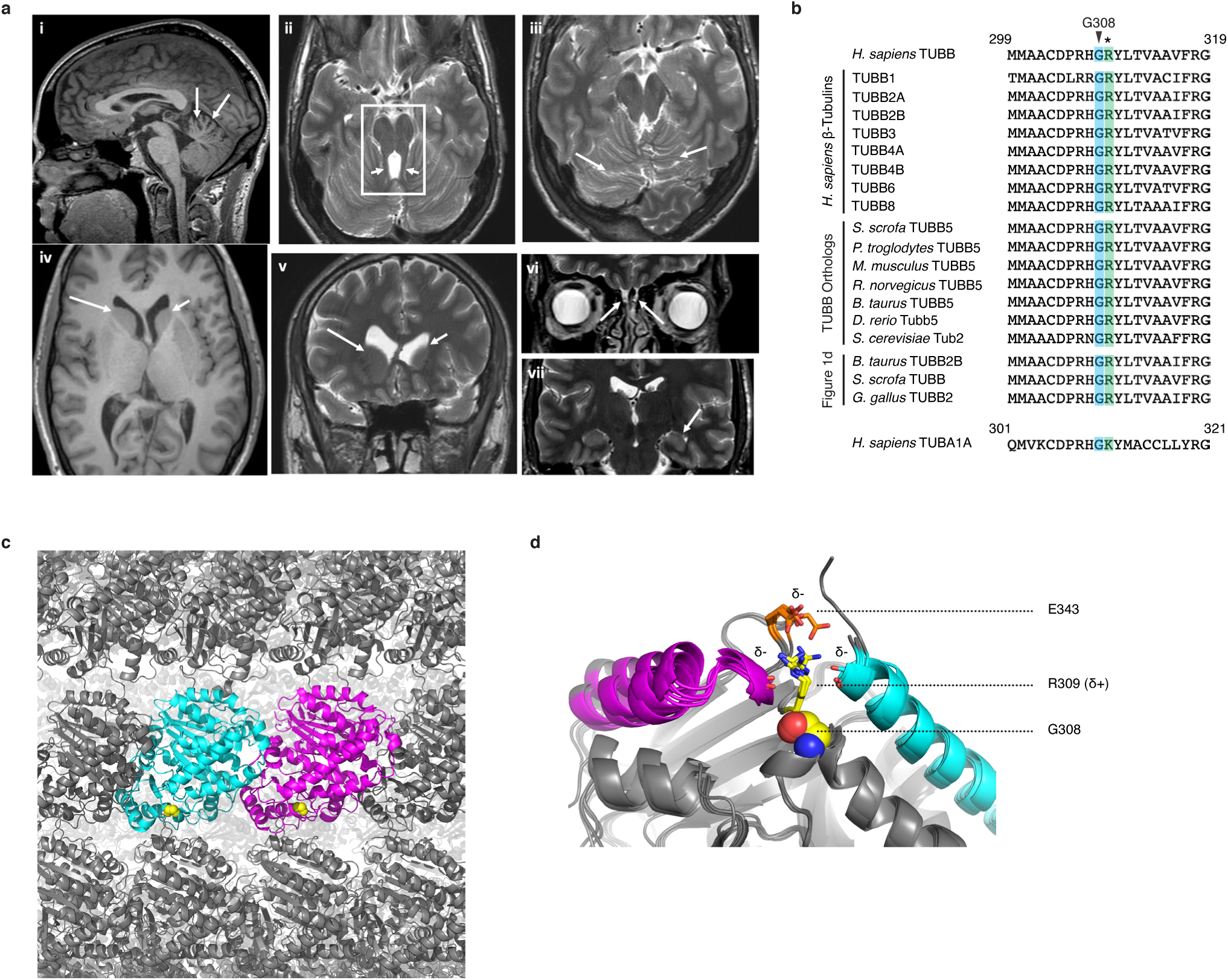
A novel *de novo* heterozygous mutation in *TUBB* was identified in a patient with atypical brain imaging features. (a) Brain magnetic resonance imaging (MRI) sections of the patient carrying the TUBB G308S mutation. [i] Sagittal T1 image showing a hypoplastic superior vermis (white arrows), which along with the axial T2 image at the level of the midbrain [ii] reveals the classic “molar tooth sign” (white box in ii) due to the appearance of the midbrain and antero-posteriorly oriented superior cerebellar peduncles (short white arrows in ii). [iii] Axial T2 image showing abnormal dysplastic superior cerebellar sulcations (white arrows). [iv] (axial T1 image) and [v] (coronal T2 image) show the enlarged right caudate nucleus head (longer white arrows) compared to the left caudate nucleus (shorter white arrows). [vi, vii] Coronal T2 images showing additional findings of hypoplastic olfactory bulbs [vi] and incomplete rotation of the left hippocampus [vii] (white arrows). (b) Protein sequence alignment of human TUBB (299-319 aa) with the other human β-tubulin isotypes, TUBB orthologs from the indicated species, the β-tubulins in Figure 1d, and human TUBA1A. G308 in TUBB and the corresponding residue in the other sequences are indicated by an arrowhead and highlighted in blue. R309 in TUBB and the corresponding residue in the other sequences are indicated by an asterisk and highlighted in green. (c) Superimposition of the human αβ tubulin dimer (PDB ID: 6i2i) onto a fully assembled microtubule from *Sus scrofa* (PDB ID: 6o2s). β-tubulin is magenta, α-tubulin is cyan. G308 is shown in space-filling spheres (*yellow*). (d) Superimposition of 5 β-tubulin structures from 4 species, including human TUBB (PDB IDs: 6i2i, 4i4t, 5nqu, 5yl2, 5ca1). G308 is shown in space-filling spheres. R309 and E343 are shown as sticks. α-helix 10 is magenta, α-helix 12 is cyan, R309 is yellow, E343 is orange, electronegative oxygen atoms are red, electropositive nitrogen atoms are blue.

Overall, the patient described here represents an atypical case of neurodevelopmental disorder showing both ciliopathy- and tubulinopathy-like brain MRI features.

### Whole genome sequencing analysis identified a presumed pathogenic variant in Tubulin Beta Class I

To identify the genetic cause of disease in the patient, we used whole genome sequencing (WGS) to analyze coding and non-coding germline variants segregating in the members of the patient’s family: the patient, the patient’s parents, and the patient’s unaffected siblings. Rare variants were assessed for pathogenicity both against the “rare multisystem ciliopathy disorders” panel (https://panelapp.genomicsengland.co.uk/panels/150) and in a gene panel-independent fashion, considering both dominant and recessive inheritance models for segregation analysis. Overall, our analysis predicted a novel *de novo* heterozygous missense variant (NM_178014.3: c.922G>A/p.G308S) in Tubulin Beta Class I (*TUBB*) as the primary pathogenic candidate, since it was the only variant meeting defined pathogenicity criteria and segregating with the disease phenotype in the family (described in **Methods**). The *TUBB* gene is associated with two disorders with an autosomal dominant mode of inheritance: a brain development condition characterized by microcephaly and structural brain abnormalities (cortical dysplasia, complex, with other brain malformations 6 - OMIM #615771)^33^, and a phenotypically complex disorder neurologically characterized by intellectual disability without gross brain malformations (circumferential skin creases Kunze type, CSC-KT - OMIM #156610)^34^. As with disease-causing variants in the other tubulin genes, all known pathogenic mutations in *TUBB* are *de novo*^5,33,34^. Notably, *TUBB* is reported to be ubiquitously expressed in adult tissues and one of the most highly expressed β-tubulin genes in the developing brain^33,35^.

Sequence alignment analysis showed that the G308 residue is conserved in all nine human β-tubulin isotypes and in all *TUBB* orthologs from yeast to primates **(Figure 1b)**. Mapping G308 on the 3D structure of the human αβ tubulin dimer and superimposing the dimer onto the structure of a fully assembled MT from *Sus scrofa* showed that G308 is exposed on the dimer surface and is not an interface residue **(Figure 1c)**. Positionally conserved in β-tubulin structures in different species, G308 lies in a sharp turn preceding β-sheet 8^36^ and is followed by an arginine residue (R309) that is similarly conserved in all human β-tubulins and in all *TUBB* orthologs from yeast to primates **(Figure 1b and 1d)**. The positive side chain of R309 occupies a structurally important position and is involved in charge interactions with the negative dipole of two α-helices (10 and 12)^36^ and the side chain of a glutamic acid residue (E343) **(Figure 1d)**. Considering the constrained position occupied by G308 and the unique flexibility of glycine residues, it is conceivable that G308 is crucial to hold R309 in the optimal position for its interactions.

### The TUBB G308S variant affects MT assembly and alters MT structure

To determine the functional impact of the patient’s TUBB G308S mutation on αβ tubulin dimer formation and incorporation into MTs, we performed a MT incorporation assay where hTERT-RPE1 cells were transiently transfected with plasmids encoding either TUBB-V5 WT or TUBB-V5 G308S, and immunostained for exogenous TUBB and endogenous α-tubulin **(Figure 2)**. In most transfected cells TUBB-V5 WT co-localized with α-tubulin and had a filamentous appearance, indicating stable incorporation into MTs **(Figure 2-i)**. Conversely, TUBB-V5 G308S had a diffuse appearance in ∼90% of transfected cells, while full or partial incorporation into MTs was observed in only ∼10% of cells **(Figure 2-ii and 2-iii**, n = 181 cells). These data suggest that TUBB G308S impairs αβ tubulin dimer formation or incorporation into MT protofilaments.

**Figure 2:**
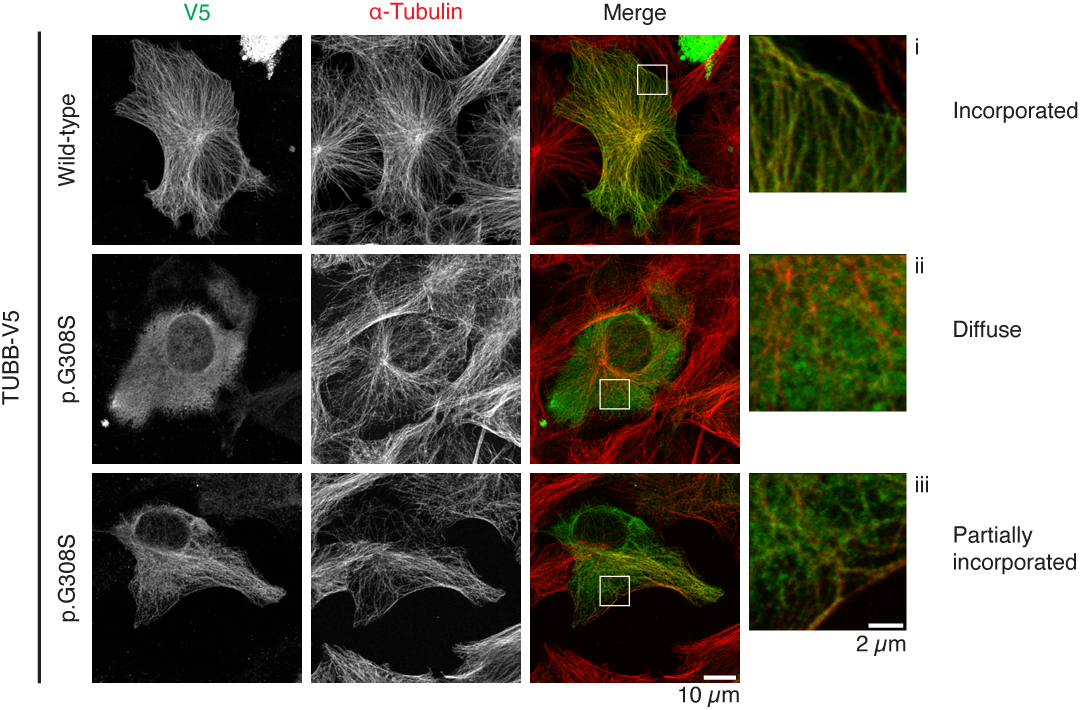
The G308S mutant TUBB protein shows reduced incorporation into microtubules. Representative images of hTERT-RPE1 cells overexpressing wild-type or G308S TUBB-V5 and immunostained for V5 (*green*) and total α-tubulin (*red*). [i, ii, iii] Magnifications of the indicated areas in the merged images (white boxes) showing complete microtubule incorporation [i], diffuse distribution [ii] and partial microtubule incorporation [iii] of TUBB-V5.

The observation that TUBB-V5 G308S showed residual MT incorporation in a small proportion of transfected cells prompted us to investigate the impact of this variant on MT function. To this end, we used prime editing^37^ to generate a TUBB G308S heterozygous cell line (TUBB^G308S^) in human diploid p53-null hTERT-RPE1 cells (TUBB^WT^). The genotype of the generated TUBB^G308S^ cell model was confirmed by Sanger sequencing **(Figure 3a)** and no editing was detected at the four most likely predicted protein-coding off-target sites **(Figure S1)**. Compared to plasmid-based studies, where the mutant protein is overexpressed and present in variable amounts across different cells, TUBB^G308S^ cells represent a more physiologically relevant system in which the allele dosage of the heterozygous patient is preserved. Semiquantitative RT-PCR for *TUBB* mRNA showed comparable expression levels in TUBB^WT^ and TUBB^G308S^ cells **(Figure 3b-i)**, where both the wild-type and the mutant alleles are transcribed at a 1:1 ratio **(Figure 3b-ii)**. Mutations in b-tubulin can result in post-translational degradation of the mutant protein^38^. However, immunoblot analysis revealed that b-tubulin levels in total cell lysates were unchanged in TUBB^G308S^ compared to TUBB^WT^ cells **(Figure S2a)**, suggesting that the G308S mutation does not alter protein stability.

**Figure 3:**
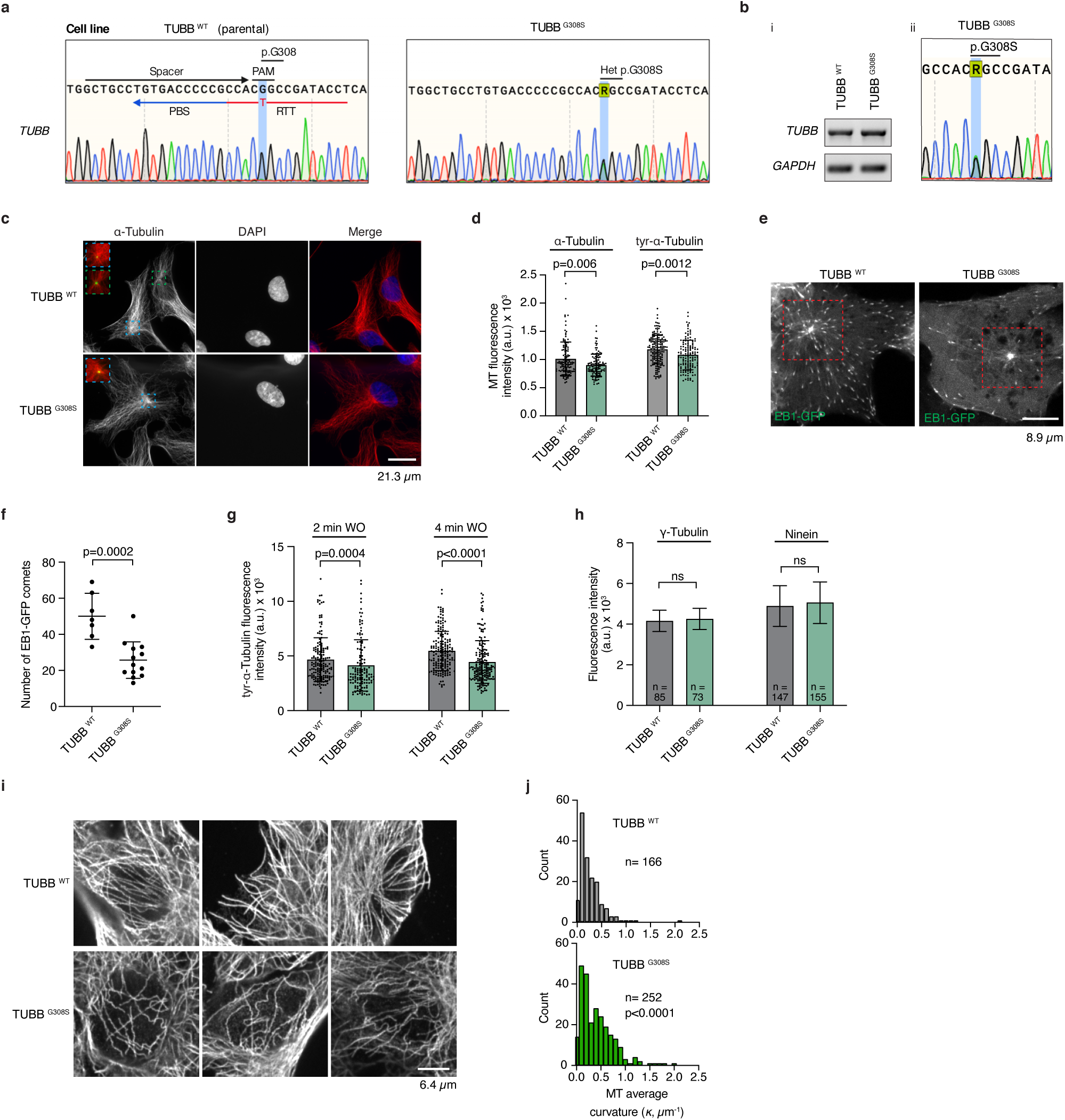
Heterozygous TUBB^G308S^ hTERT-RPE1 cells nucleate fewer microtubules that show excessive curvature. (a) Schematic representation of the prime editing guide RNA (pegRNA) used for editing the *TUBB* locus and Sanger sequencing confirmation of the generated TUBB^G308S^ cell model in hTERT-RPE1 cells. (b) [i] Semiquantitative RT-PCR analysis of *TUBB* mRNA in parental TUBB^WT^ cells and the TUBB^G308S^ cell model. [ii] Sanger sequencing electropherogram of *TUBB* cDNA from TUBB^G308S^ cells (c) Representative images of the MT network in cells fixed and immunostained for α-tubulin (*red*), DNA (*blue*), and ninein (*green*). (d) Scatter plot of MT mean fluorescence intensity measured around the centrosome region in cells fixed and immunostained for α-tubulin (n ≥ 119 cells) and tyrosinated-α-tubulin (n ≥ 134 cells). P-values were calculated using the Mann U Whitney test and unpaired two-tailed Student’s t-test, respectively. (e) Confocal projections of EB1-GFP in TUBB^WT^ and TUBB^G308S^ cells. (f) Scatter plot of the number of EB1-GFP comets analyzed in a fixed-area box at the centrosome (red box in e), (n = 7 cells for TUBB^WT^ and n = 13 cells for TUBB^G308S^). P-value calculated using an unpaired t-test. (g) Scatter plot of tyrosinated-α-tubulin mean fluorescence signal emanating from centrosomes after nocodazole washout (WO) and MT regrowth for 2 min (n ≥ 128 cells) and 4min (n ≥ 177 cells). P-value calculated using Mann U Whitney test. (h) Bar graph of mean fluorescence intensity of γ-tubulin and ninein at the centrosome. P-value calculated using an unpaired t-test. (i) Examples of single confocal sections of MTs from TUBB^WT^ and TUBB^G308S^ cells. (j) Histogram of MT average curvature in TUBB^WT^ and TUBB^G308S^ cells. MTs from 27 and 47 cells were analyzed for TUBB^WT^ and TUBB^G308S^, respectively. P-value calculated using Mann U Whitney test. Data are represented as mean ± standard deviation. ns = not statistically significant (p > 0.05).

To understand how the TUBB G308S mutation impacts MT organization and dynamics, we first examined the entire MT array in TUBB^WT^ and TUBB^G308S^ cells by immunofluorescence. TUBB^G308S^ cells showed overall dimmer α-tubulin intensity than TUBB^WT^ cells **(Figure 3c)**. Indeed, the mean α-tubulin fluorescence intensity around the ninein-marked centrosome was significantly reduced in TUBB^G308S^ cells compared to TUBB^WT^ cells **(Figure 3d)**. A similar reduction in fluorescence intensity was observed when staining for newly formed MTs marked by tyrosinated α-tubulin **(Figure 3d)**. This finding suggests that MT nucleation or early polymerization is impaired in TUBB^G308S^ cells. To better visualize MT assembly in TUBB^G308S^ cells, we performed imaging of live cells transiently transfected with the MT plus-end binding protein EB1-GFP to monitor MT nucleation and growth **(Figure 3e)**. We quantified the number of EB1-GFP comets in cells expressing weak and medium EB1-GFP levels. Intriguingly, we found a significant decrease in the number of EB1-GFP comets emanating from the centrosome in TUBB^G308S^ cells compared to TUBB^WT^ cells **(Figure 3e and 3f)**. To complement these observations, we assessed MT regrowth after nocodazole treatment and washout in TUBB^G308S^ and TUBB^WT^ cells. Quantifying levels of tyrosinated α-tubulin after 2 or 4 minutes of recovery showed lower fluorescence intensity in TUBB^G308S^ cells compared to TUBB^WT^ cells **(Figure 3g)**, suggesting that fewer MTs are nucleated and assembled in mutant tubulin-expressing cells. To determine if this was from a defect in the MT organizing centre (MTOC), we examined the centrosomal recruitment of γ−tubulin and ninein, two proteins required for MT polymerization from the centrosome^39^. The centrosomal levels of both γ−tubulin and ninein were similar in TUBB^G308S^ cells compared to control cells **(Figure 3h)**, suggesting that TUBB^G308S^ cells have intact MTOC structures but with lower MT nucleating capacity.

We also noticed, during imaging of the MT array, that MT architecture was different in TUBB^G308S^ cells, where wavy and buckled MTs were frequently observed **(Figure 3i)**. To analyze this further, we acquired thin confocal sections of individual MT filaments and measured their curvature. Analysis of MT curvature (detailed in **Methods**) showed that the distribution of the average MT curvature in TUBB^G308S^ cells deviated significantly from that of TUBB^WT^ cells by being wider and shifted to the right **(Figure 3j)**. To exclude potential fixation artifacts, we also examined MT curvature in live cells by tracking EB1-GFP trajectories in TUBB^G308S^ cells compared to TUBB^WT^ cells. MT curvature was measured after tracing maximum intensity confocal projections of EB1-GFP in time-lapse series **(Figure S2b)**. Consistent with our immunostaining results, the EB1-GFP curvature distribution in mutant TUBB^G308S^ cells was significantly different from that of TUBB^WT^ cells **(Figure S2c)**, suggesting that newly formed MTs in TUBB^G308S^ cells are subjected to structural alterations caused by the mutation. We then hypothesized that excessive MT curvature could affect the ability of MT-associated proteins to optimally interact with MTs in TUBB^G308S^ cells. Indeed, we found a significant decrease in the mean fluorescence of EB1-GFP comets in TUBB^G308S^ cells compared to TUBB^WT^ cells **(Figure S2d)**. Collectively, these results show that the TUBB G308S mutation affects MT assembly and overall MT organization.

### The TUBB G308S variant causes a primary cilia defect

The ciliopathy-like features in the patient’s brain MRI prompted us to investigate the potential effect of the newly identified TUBB G308S variant on primary cilia. To this end, we first sought to characterize cilia-related phenotypes in primary skin fibroblasts from the patient and an age- and sex-matched healthy control **(Figure 4a)**. To validate primary skin fibroblasts as a suitable cell type for this purpose, we verified that *TUBB* is expressed. Semiquantitative transcript analysis by RT-PCR revealed that *TUBB* is highly expressed in skin fibroblasts, and at similar levels in control and patient cells **(Figure 4b-i)**. Moreover, both the wild-type and the mutant alleles are transcribed in patient cells, at a 1:1 ratio **(Figure 4b-ii)**. To study primary cilia in cell culture, ciliation is commonly induced by promoting cell cycle exit through serum starvation. Immunostaining of primary cilia in serum-starved quiescent fibroblasts **(Figure 4c)** revealed that patient cells exhibited a significant decrease (∼30%) in ciliation frequency compared to control cells **(Figure 4c and 4d)**, while showing normal cilium length **(Figure 4e)**. This finding supports a role for ciliation defects in the patient’s clinical presentation and provides a cellular phenotype for subsequent *in vitro* studies.

**Figure 4:**
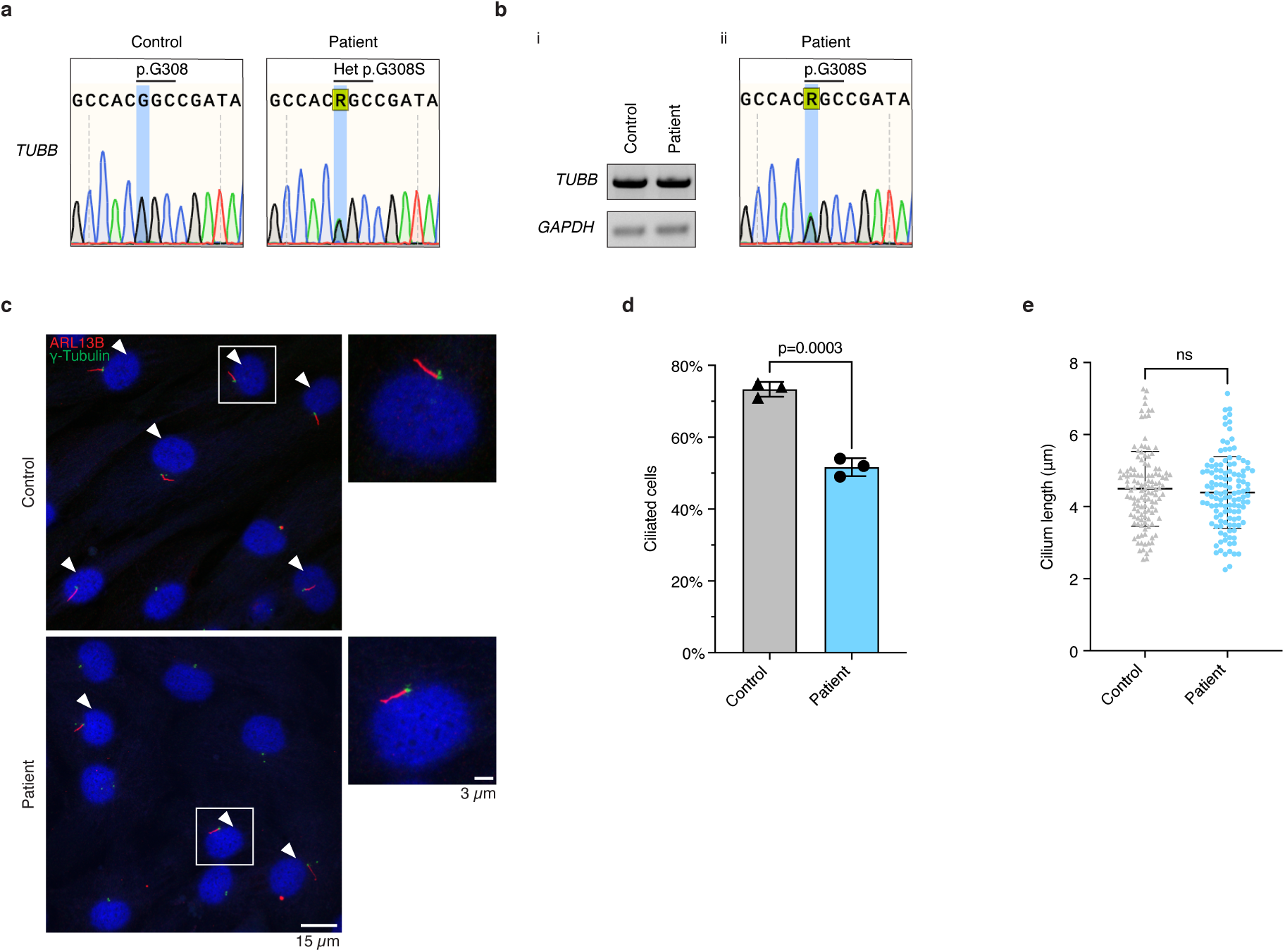
Patient fibroblasts show reduced ciliation and normal cilium length. (a) Sanger sequencing electropherograms of the *TUBB* locus in primary skin fibroblasts from the patient and an age- and sex-matched healthy control subject. (b) [i] Semiquantitative RT-PCR analysis of *TUBB* mRNA in control and patient fibroblasts. [ii] Sanger sequencing electropherogram of *TUBB* cDNA from patient fibroblasts. (c) Representative images of control and patient fibroblasts fixed after 72 h serum withdrawal and immunostained for the ciliary marker ARL13B (*red*) and the centrosomal marker γ-tubulin (*green*). White arrowheads indicate ciliated cells. Right panels show magnifications of the indicated cells (white boxes) (d) Percentage of cells showing ARL13B-positive cilium in control and patient fibroblasts after 72 h serum withdrawal. Data from three experiments, each with two technical replicates (n ≥ 400 cells) are shown. P-value was calculated using an unpaired t-test. (e) Scatter plot of the length of ARL13B-positive cilia in control (n = 123 cilia) and patient (n = 122 cilia) fibroblasts after 72 h serum withdrawal. P-value was calculated using an unpaired t-test. Data are represented as mean ± standard deviation. ns = not statistically significant (p > 0.05).

To associate the ciliation phenotype observed in patient cells with the TUBB G308S variant, the effect of the mutation must be studied in a context that is different from the patient’s genetic background, and isogenic between control and mutant cells. Human hTERT-RPE1 cells are a common model to study primary cilia *in vitro* due to efficient ciliation and the functional properties of their cilium^40^. Therefore, we assessed ciliation in the TUBB^G308S^ cell model compared to the isogenic TUBB^WT^ cell line. Importantly, immunostaining of cilia in serum-starved cells **(Figure 5a)** revealed that the TUBB^G308S^ cell model exhibited a significant decrease (∼35%) in ciliation frequency compared to control cells **(Figure 5a and 5b)**. Notably, the magnitude of this reduction was similar to that observed in patient fibroblasts. The cell model accurately recapitulates the ciliation phenotype observed in patient cells, showing that the TUBB G308S mutation impairs ciliation and reinforcing the hypothesis that it is causative of the patient’s clinical phenotype.

**Figure 5:**
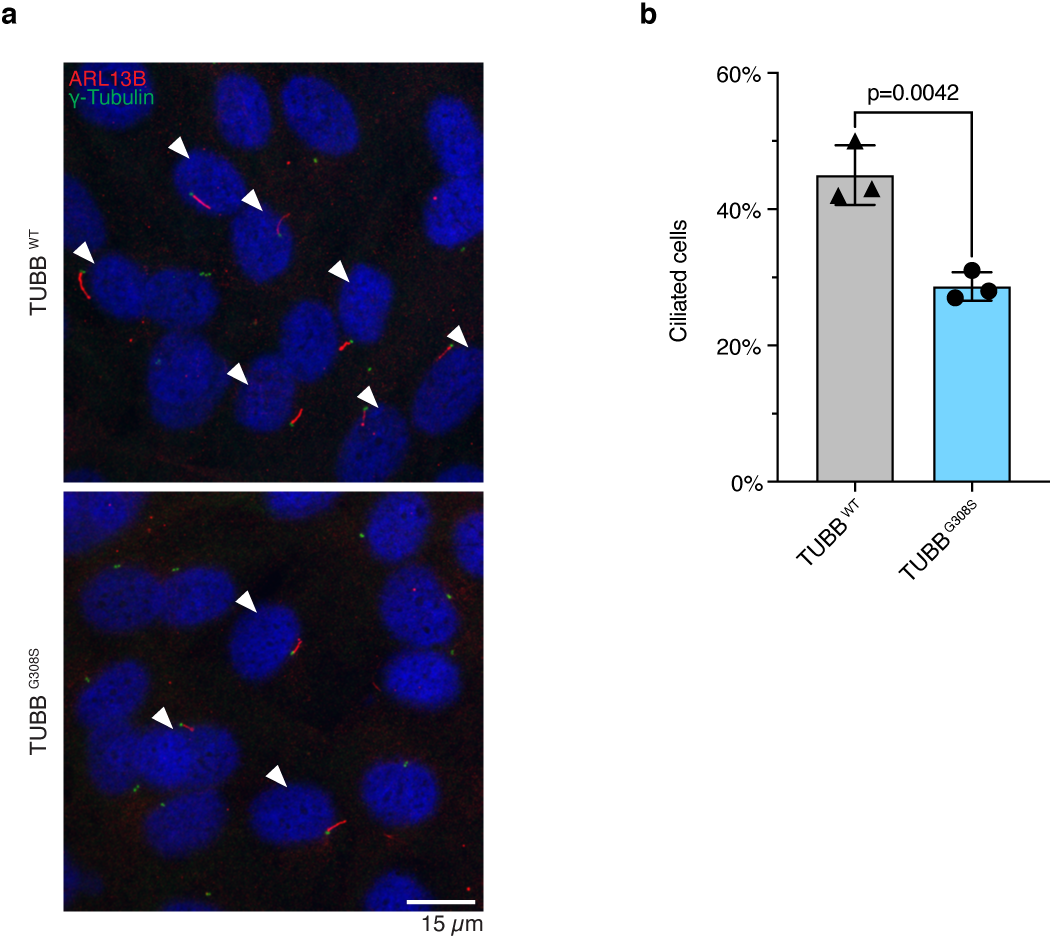
Heterozygous TUBB^G308S^ hTERT-RPE1 cells show reduced ciliation. (a) Representative images of TUBB^WT^ and TUBB^G308S^ cells fixed after 48 h serum withdrawal and immunostained for the ciliary marker ARL13B (*red*) and the centrosomal marker γ-tubulin (*green*). White arrowheads indicate ciliated cells. (b) Percentage of cells showing ARL13B-positive cilium in TUBB^WT^ and TUBB^G308S^ cells after 48 h serum withdrawal. Data from three experiments, each with four technical replicates (n = 600 cells) are represented as mean ± standard deviation. P-value was calculated using an unpaired t-test.

### The TUBB G308S variant impairs primary cilia formation and centrosome migration

We next sought to understand the molecular mechanism by which the TUBB G308S mutation impairs ciliation. Since the primary cilium is assembled in the G0/G1 phase of the cell cycle^23^ and MT function and organization are also closely interconnected with cell cycle progression^41,42^, we first wondered if the TUBB G308S mutation affects ciliation by altering cell cycle regulation. To test this, we investigated whether TUBB^G308S^ cells are able to exit the cell cycle upon serum starvation. Cell cycle analysis by flow cytometry revealed that, despite cycling slower than TUBB^WT^ cells, TUBB^G308S^ cells responded to serum starvation **(Figure S3a and S3b)**. Most importantly, TUBB^G308S^ cells exited the cell cycle as effectively as TUBB^WT^ cells and no difference in the proportion of quiescent cells was observed following serum starvation in the conditions where the ciliation defect was apparent **(Figure S3c)**. This finding rules out the hypothesis that the observed ciliation phenotype is an indirect consequence of a primary cell cycle defect.

We next investigated whether the ciliogenesis pathway is compromised in mutant cells. hTERT-RPE1 cells rely on the intracellular pathway of ciliogenesis^25,26^, which is the same pathway used by neurons and neuronal progenitors^25–27^, the relevant cell types in neurodevelopmental disorders. In intracellular ciliogenesis, dynamic MTs play a critical role in trafficking PCVs to the centrosome and in the subsequent migration of the forming cilium to the plasma membrane^28,30,32^. In hTERT-RPE1 cells, PCVs trafficking occurs within the first few hours of serum withdrawal, when axoneme elongation begins^28,30^. To test whether the TUBB G308S mutation affects early stages of ciliogenesis, we quantified the presence of detectable cilia in in TUBB^WT^ and TUBB^G308S^ cells after 2, 4 or 6 hours of serum withdrawal. We found that TUBB^G308S^ cells showed significantly lower ciliation frequencies than control cells from as early as 4 hours after serum withdrawal **(Figure 6a)**, supporting the impact of the TUBB G308S mutation on the early steps of cilia formation.

**Figure 6:**
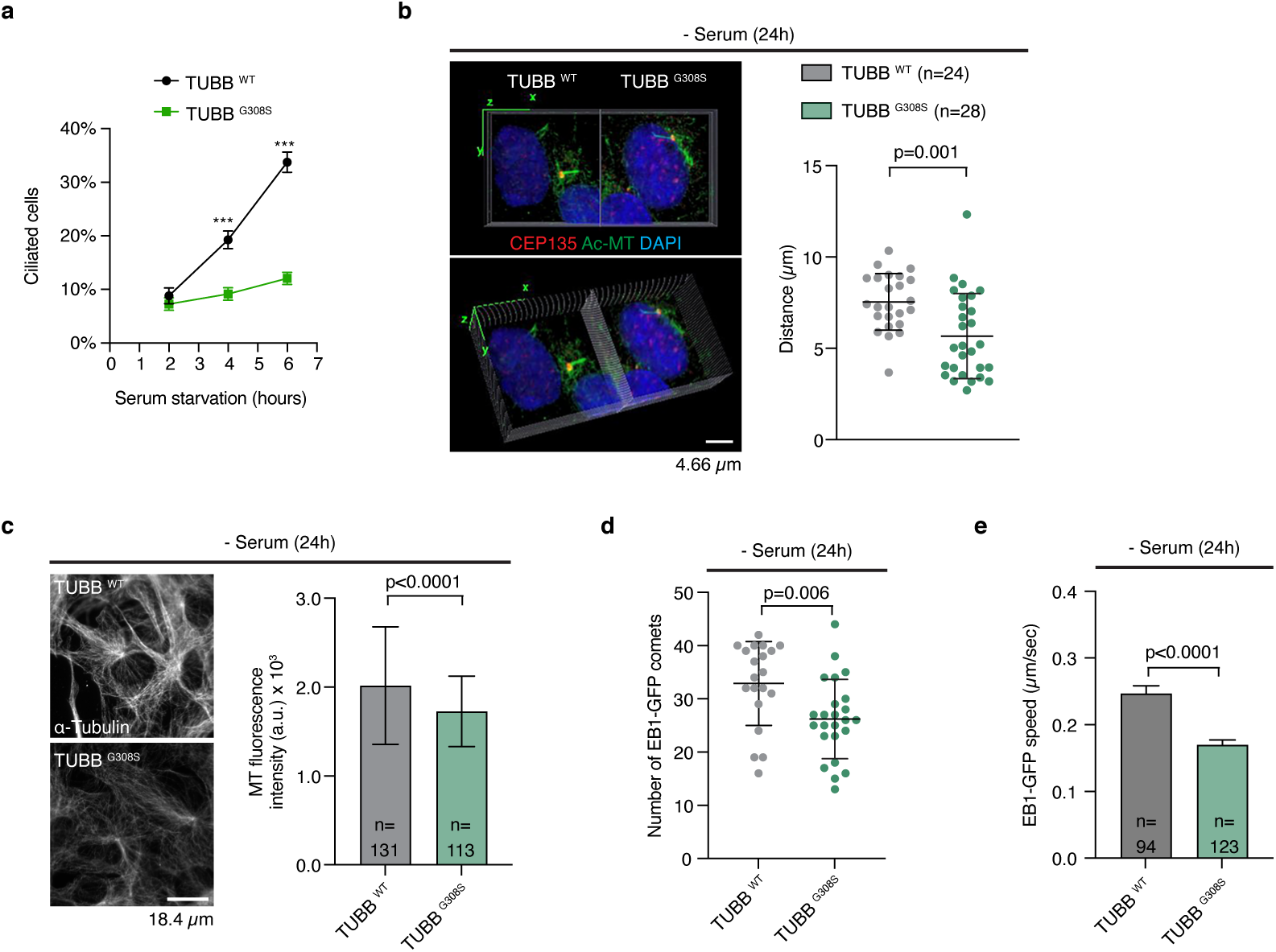
Heterozygous TUBB^G308S^ hTERT-RPE1 cells show defective primary cilia formation and migration. (a) Percentage of cells showing acetylated-α-tubulin-positive cilium in TUBB^WT^ and TUBB^G308S^ cells after serum withdrawal for the indicated time points (n ≥ 374; *** indicate p < 0.0001 by Šídák’s multiple comparisons test) (b) Volume reconstruction of confocal z-stack images of primary cilia in fixed TUBB^WT^ and TUBB^G308S^ cells after 24 h serum withdrawal. Cells were immunostained for the centrosomal marker CEP135 (*red*), acetylated-α-tubulin (*green*) and DNA (*blue*) to label the nucleus. Scatter plot of the migration distance of ciliated centrosomes. Migration was measured on the XZ plane projection as the distance between CEP135 and the apex of the nucleus. P-value was calculated using an unpaired t-test. (c) Representative images of the MT network in TUBB^WT^ and TUBB^G308S^ cells fixed and immunostained for α-tubulin following 24h serum withdrawal. Bar graph of MT mean fluorescence intensity measured around the centrosome region. P-value was calculated using an unpaired t-test. (d) Scatter plot of the number of EB1-GFP comets following 24h serum withdrawal. The number of EB1-GFP comets was analyzed in a fixed-area box at the centrosome (n = 20 cells for TUBB^WT^ and 24 cells for TUBB^G308S^). P-value was calculated using an unpaired t-test. (e) Bar graph of EB1-GFP comet speed following 24 h serum withdrawal. P-value was calculated using Mann U Whitney test. Data are represented as mean ± standard deviation (b, c, d) or ± standard error of the mean (a, e)

MT pushing forces, generated by growing MTs, are proposed to apically move the centrosome/forming cilium complex towards the plasma membrane^32^. Therefore, we wondered whether the MT nucleation defects and MT deformations observed in TUBB^G308S^ cells **(Figure 3)** affect the MT subpopulations contributing to the migration of the primary cilium to the plasma membrane. To test this hypothesis, we induced primary cilia formation by serum starving cells for 24 hours before immunostaining for primary cilia and the centrosomal protein CEP135 **(Figure 6b)**. Confocal images encompassing the entire thickness of cells were acquired **(Figure 6b)** and converted to XZ projections to measure the relative distance between the ciliated centrosome and the nuclear apex. We found that ciliated centrosomes in TUBB^G308S^ cells traveled shorter distances compared to their counterpart in control cells **(Figure 6b)**. Similar results were obtained when measuring the spatial proximity of ciliated centrosomes to the nearest nucleus margin **(Figure S4a and S4b)**.

To investigate the underlying molecular mechanism of the reduced primary cilia formation and migration, we analyzed the assembly and structure of MTs in serum-starved cells. Immunofluorescence analysis of total MTs in TUBB^G308S^ cells showed a reduced MT density compared to control cells also in serum-starved conditions **(Figure 6c)**. We also analyzed MT nucleation through EB1-GFP expression in control and mutant cells under serum-starvation. TUBB^G308S^ cells showed significantly less MT nucleation events compared to TUBB^WT^ cells **(Figure 6d and Supplementary video 1)**, as well as reduced EB1-GFP accumulation at MT plus-ends **(Figure S4c)**. Since MT pushing forces are involved in primary cilia migration, we also examined the speed of EB1-GFP comets and found MTs in TUBB^G308S^ cells had reduced growth speed compared to MTs in control cells **(Figure 6e)**.

Collectively, our results suggest that the TUBB G308S mutation impairs primary cilia formation and migration, due to the critical roles of MT growth, dynamics and organization in ciliogenesis.

### *TUBB* gene haploinsufficiency does not affect ciliation

Next, we intended to investigate the association between mutations in the *TUBB* gene and defective ciliation beyond the patient’s G308S variant. While all known pathogenic mutations in tubulin genes are dominant^4^, both gain-of-function and haploinsufficiency pathogenic mechanisms have been proposed^33,43–45^.

To evaluate the contribution of *TUBB* haploinsufficiency to the ciliation phenotype, a *TUBB* haploid cell model was generated in hTERT-RPE1 cells using CRISPR/Cas9^46^. Haploidization of the *TUBB* locus was achieved by delivering a pair of guide RNAs (gRNAs) that span the *TUBB* gene from intron 1 to the downstream intergenic region **(Figure 7a)**, with the intronic gRNA being specific to a naturally occurring heterozygous SNP in hTERT-RPE1 cells **(Figure 7a-i)**. This genome editing strategy resulted in *TUBB* truncation after exon 1 (19 amino acids) on the targeted allele, leaving the *TUBB* gene intact on the non-targeted allele **(Figure 7a-ii and 7a-iii)**. Copy number analysis by digital droplet PCR (ddPCR) confirmed the successful haploidization of the *TUBB* locus **(Figure 7b)**. Gene expression analysis by quantitative RT-PCR (qPCR) showed a ∼30% decrease in *TUBB* mRNA levels in the haploidized cell line compared to wild-type cells, suggesting partial compensation for the deleted allele by *TUBB* haploid cells **(Figure 7c)**. Importantly, immunostaining of cilia in serum-starved cells **(Figure 7d)** revealed no difference in ciliation frequency between *TUBB* haploid and wild-type cells **(Figure 7d and 7e)**. This finding indicates that *TUBB* haploinsufficiency does not affect ciliation in hTERT-RPE1 cells.

**Figure 7:**
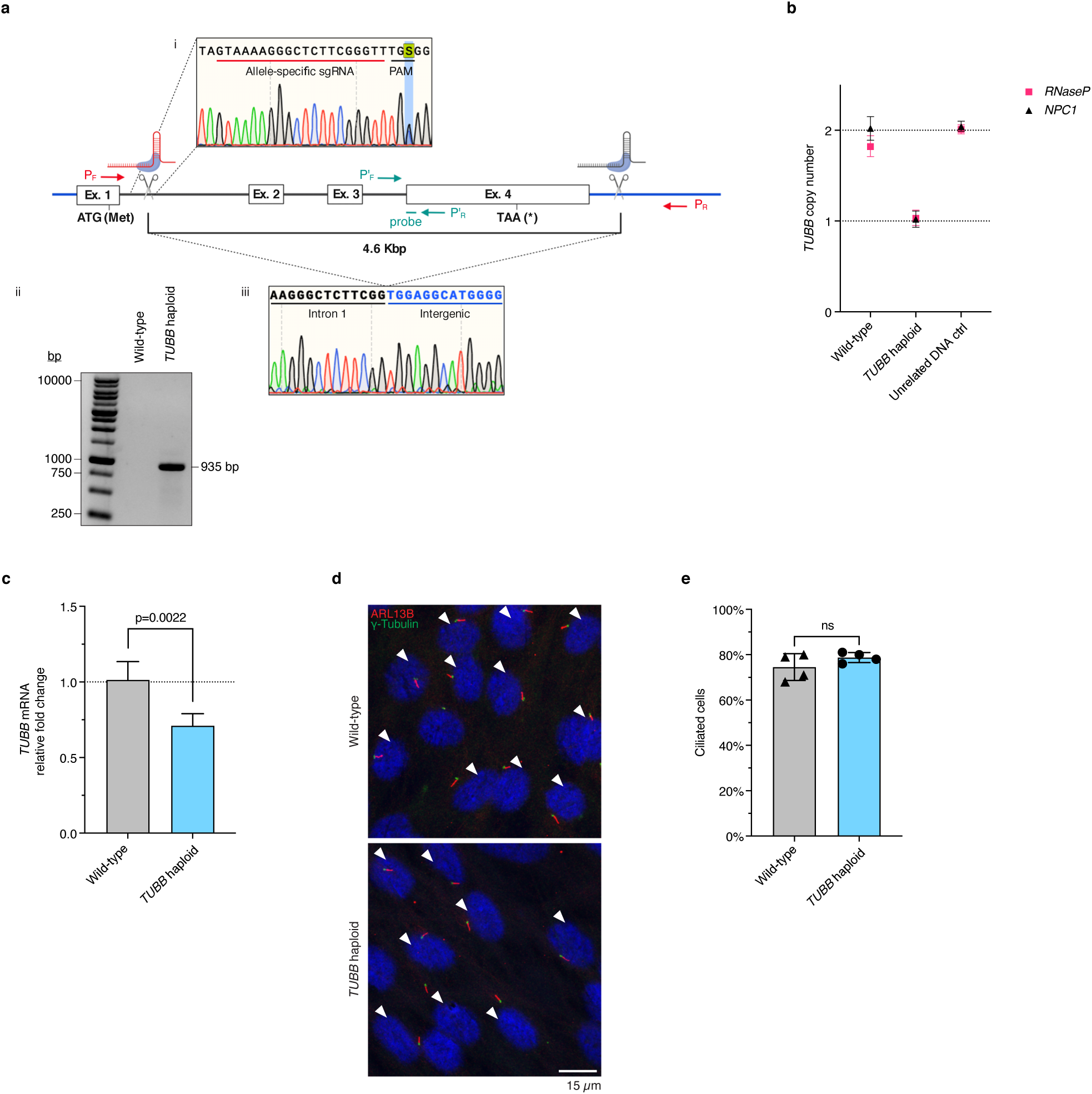
*TUBB* haploidized hTERT-RPE1 cells show normal ciliation. (a) Schematic representation of the genome editing strategy used to haploidize the *TUBB* locus. [i] Sanger sequencing electropherogram of the target heterozygous site in wild-type hTERT-RPE1 cells and representation of the guide RNA used for allele-specific editing. [ii] PCR amplification spanning the deleted region in wild-type and *TUBB* haploidized cells using primer pair P_F_-P_R_ (*red*). PCR conditions did not allow to amplify the intact fragment from wild-type cells because it is too large. [iii] Sanger sequencing electropherogram of the deletion junction in *TUBB* haploidized cells (b) Plot of *TUBB* copy number relative to two reference genes (*RNaseP* and *NPC1*) from digital droplet PCR (ddPCR) analysis in wild-type and *TUBB* haploidized hTERT-RPE1 cells, and blood-derived human DNA as an unrelated diploid control. The assay was performed using primer pair P’_F_-P’_R_ and a probe located within the deleted region (*green* in a). Error bars represent the 95% confidence interval limits. (c) *TUBB* mRNA expression levels normalized to *GAPDH* and plotted relatively to wild-type hTERT-RPE1 cells from quantitative PCR (qPCR) analysis. Data from three experiments are represented as mean ± standard error of the mean. P-value was calculated using a paired t-test on delta Ct values. (d) Representative images of wild-type and *TUBB* haploidized hTERT-RPE1 cells fixed after 48 h serum withdrawal and immunostained for the ciliary marker ARL13B (*red*) and the centrosomal marker γ-tubulin (*green*). White arrowheads indicate ciliated cells. (e) Percentage of cells showing ARL13B-positive cilium in wild-type and *TUBB* haploidized hTERT-RPE1 cells after 48 h serum withdrawal. Data from four experiments, each with four technical replicates (n = 600 cells) are represented as mean ± standard deviation. P-value was calculated using an unpaired t-test. ns = not statistically significant (p > 0.05).

### More clinically relevant dominant mutations in *TUBB* affect ciliation *in vitro*

We next sought to investigate the effect of other clinically relevant mutations in *TUBB* on primary cilia. In particular, we considered all the sixteen non-synonymous single nucleotide polymorphism (SNP) variants present in the ClinVar database in April 2020.

Since mutations in the *TUBB* gene do not affect ciliation by haploinsufficiency **(Figure 7e)**, we reasoned it should be possible to elicit their gain-of-function phenotype and test their effect in an overexpression system. To this end, hTERT-RPE1 cells were transiently transfected with either TUBB-V5 WT or TUBB-V5 bearing a specific variant, serum starved the following day for 24 hours to induce ciliogenesis, and the percentage of ciliated cells was assessed in transfected (V5 positive) and non-transfected (V5 negative) cells **(Figure 8a and 8b)**. Importantly, overexpression of the patient’s G308S variant caused a dramatic decrease (∼90%) in ciliation frequency compared to cells overexpressing the wild-type protein **(Figure 8b)**. This finding confirms that the TUBB G308S mutation impairs ciliation, and validates our overexpression strategy for variant testing. Notably, the magnitude of the ciliation defect was substantially higher upon overexpression than it was in patient fibroblasts or in the TUBB^G308S^ cell model **(Figure 4d and 5b)**, suggesting a dose-dependent gain-of-function pathogenic mechanism for the G308S variant.

**Figure 8:**
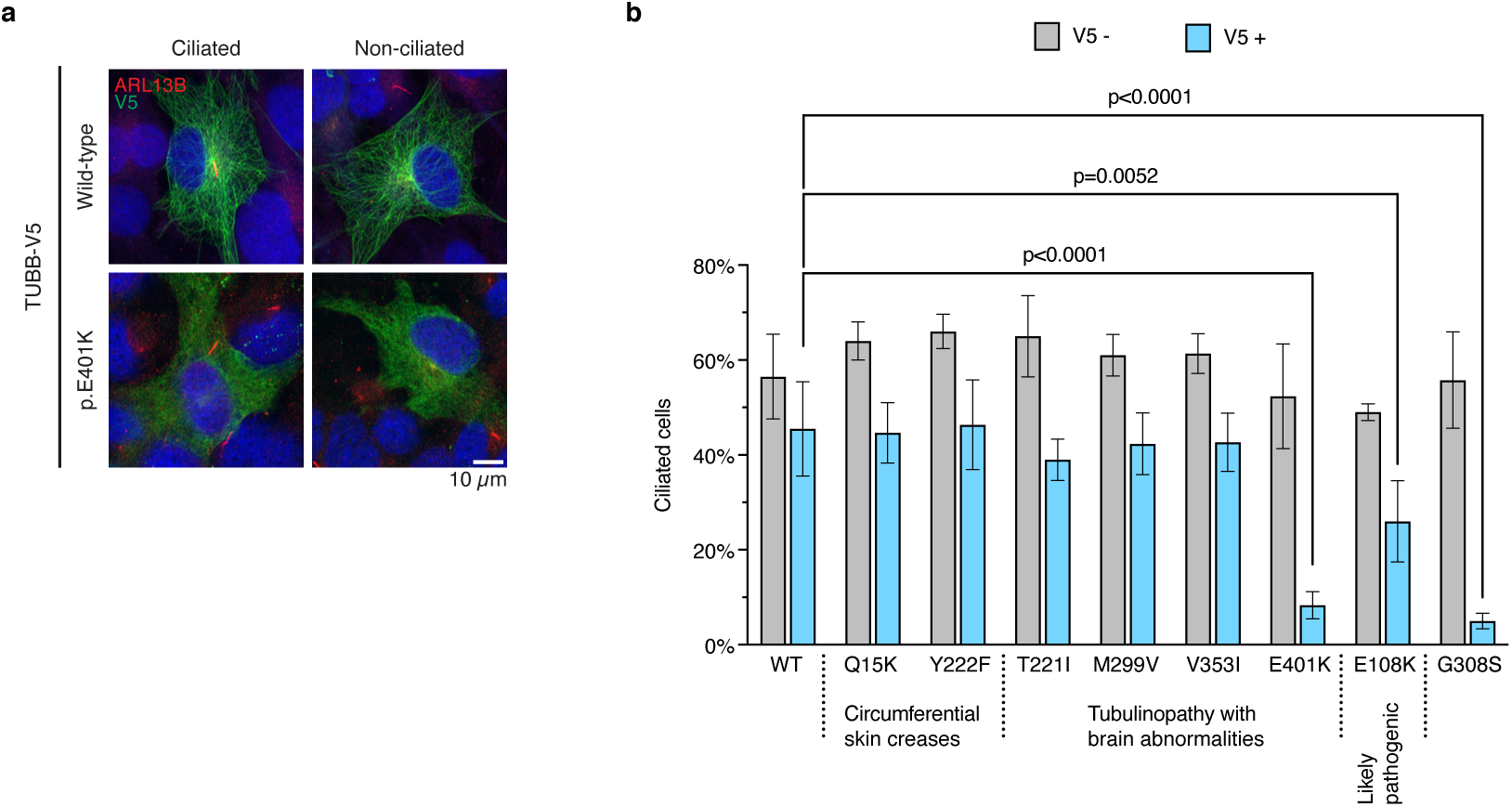
A subset of clinically relevant dominant mutations in TUBB affects ciliation in hTERT-RPE1 cells. (a) Representative images of ciliated and non-ciliated hTERT-RPE1 cells overexpressing wild-type or mutant (E401K) TUBB-V5, fixed after 24 h serum withdrawal and immunostained for V5 (*green*) and the ciliary marker ARL13B (*red*). (b) Ciliation frequency of cells overexpressing wild-type or mutant (Q15K, Y222F, T221I, M299V, V353I, E401K, E108K, G308S) TUBB-V5 after 24 h serum withdrawal. Percentage of cells showing ARL13B-positive cilium was measured in the transfected (V5-positive) and non-transfected (V5-negative) cell populations. For each mutation, the associated condition or the clinical significance from the ClinVar database is shown. Pooled data from at least three experiments (n ≥ 200 cells for V5-positive population, n = 400 cells for V5-negative population) are represented as mean ± standard deviation. P-value was calculated using one-way ANOVA followed by Dunnett’s multiple comparisons test between the wild-type control sample and each mutant, in both the transfected and non-transfected cell populations. Non-statistically significant (p > 0.05) comparisons are not shown.

Previously reported pathogenic mutations in *TUBB* are associated with CSC-KT (OMIM #156610)^34^, which do not cause gross brain malformations, and with a specific tubulinopathy characterized by structural brain abnormalities (OMIM #615771)^33^. As we expected, overexpression of the CSC-KT causing variants (Q15K and Y222F) did not cause any ciliation defect. However, overexpression of the tubulinopathy associated variant E401K — but not of T221I, M299V or V353I — caused a substantial decrease (∼80%) in ciliation frequency compared to cells overexpressing the wild-type protein **(Figure 8b)**. With regard to the variants with undetermined clinical significance in ClinVar, all ten were preliminarily tested in single-replicate experiments. No effect on ciliation was found for the variants classified as “likely benign” or “of uncertain significance”, while the “likely pathogenic” variant E108K showed reduced ciliation and was prioritized for replicate studies **(Figure S5a)**. Overexpression of the E108K variant was confirmed to affect ciliation, although in a milder way compared to G308S or E401K **(Figure 8b)**.

In the same experiment, the ability of each TUBB variant to incorporate into MTs was assessed based on the filamentous, diffuse or partially diffuse appearance of the V5-tagged exogenous TUBB protein **(Figure S5b and S5c)**. Importantly, our results were concordant with the existing literature for all the variants on which incorporation studies had been previously performed (Q15K, Y222F, M299V, V353I, E401K)^33,34^. Interestingly, a correlation was found between the ability of each TUBB variant to incorporate into MTs and its effect on ciliation. Incorporation defects were observed only for the three variants that also affected ciliation: G308S, E401K and E108K. As with ciliation, G308S and E401K showed a strong incorporation defect while E108K displayed a milder phenotype **(Figure S5c)**.

These findings show that some clinically relevant dominant mutations in *TUBB* affect ciliation *in vitro*, putting forward ciliation defects as a contributing pathogenic factor in a subset of tubulinopathy patients.

## DISCUSSION

Despite MTs being fundamental to primary cilia formation and function, no association between mutations in tubulin genes and primary ciliopathies has been found to date, and tubulinopathies and neurodevelopmental ciliopathies are considered as two independent groups of disorders. Here we present evidence suggesting that defective ciliogenesis is a contributing pathogenic factor in at least a subset of tubulinopathy patients. We show that three dominant mutations in a β-tubulin gene (*TUBB*), two pathogenic variants from patients with structural brain malformations (G308S and E401K) and one likely pathogenic variant from a patient whose clinical information is not publicly available (E108K), affect ciliation *in vitro*. We identified the TUBB G308S variant in a patient with brain abnormalities reminiscent of both JS and tubulinopathies, and validated it as the *bona fide* disease-causing mutation by showing that it affects MT dynamics and structure, and impairs ciliogenesis *in vitro*. We also showed that *TUBB* haploinsufficiency does not impair ciliation, and identified E401K and E108K in a screen for gain-of-function dominant ClinVar mutations in *TUBB* affecting brain development and ciliation.

Consistent with our finding that heterozygous loss of *TUBB* does not affect ciliation, the three variants all act in a gain-of-function fashion and impair ciliation upon overexpression in wild-type cells **(Figure 8b)**. However, the molecular mechanism by which each variant affects ciliation could be different. The novel G308S variant identified in this work shows reduced incorporation into cellular MTs in overexpression studies, affects MT nucleation and alters MT architecture. From a structural point of view, G308 seems key in positioning R309 for charge interactions that are important for TUBB tertiary structure. Interestingly, the same pattern is also present in all human α-tubulins, where G308 is conserved and R309 is replaced by a lysine residue that provides similar electrostatic properties (TUBA1A is shown as an example in **Figure 1b**). Considering the structural role of G308 and the unique flexibility of glycine residues, we hypothesized that the G308S mutation could have a subtle effect on the TUBB fold, where the slightly bulkier serine does not prevent TUBB from folding but makes the fold less stable. Consistent with this idea, *in silico* protein stability analysis predicted G308S to be among the most destabilizing glycine to serine substitutions in β-tubulins across different species **(Figure S6)**. In a plausible scenario, the mutant protein forms unstably folded αβ tubulin dimers which mostly fail to stably incorporate into MTs. Since TUBB is one of the most abundant β-tubulin isotypes, this would result in a net nucleation defect because MT assembly is a cooperative process where higher stabilization energy is required for the initial nucleation stage compared to the later elongation phase^47^. For this reason, stochastic incorporation of mutant αβ dimers could instead be tolerated during elongation, resulting in the observed curved MTs. Moreover, the tendency of mutant αβ dimers to form curved MTs likely affects nucleation itself, as it was recently shown that curved tubulin oligomers reduce nucleation efficiency by hindering lateral interactions between growing protofilaments and destabilizing early nucleation intermediates^48,49^. Similar abnormalities in MT organization and dynamics persist upon induction of ciliogenesis in serum-starved TUBB^G308S^ cells. Impaired ciliation is apparent at early time points after serum withdrawal, suggestive of defective primary cilia formation rather than reduced stability of the formed cilia. In the early stages of ciliogenesis, dynein-mediated MT transport is crucial to recruit PCVs to the mother centriole^28,30^. In mammalian cells, dynein is mainly targeted to the plus-end of MTs via a cascade of protein interactions ultimately dependent on EB1 recruitment onto MTs^50^. In serum-starved TUBB^G308S^ cells, we observed a significant reduction of EB1 accumulation on MTs. Reduced recruitment of EB1 was recently associated to different types of structural alterations of the MT plus-end, including those induced by a dominant missense mutation in the β-tubulin gene *TUBB2B* causing brain malformations^51,52^. Furthermore, variations in the overall MT structure such as bending and buckling have been shown to contribute to motor-mediated MT transport and influence vesicle motion^53^. Overall, we hypothesize that the nucleation of fewer MTs and the structural abnormalities of the existing ones affect dynein-mediated transport of PCVs to the mother centriole resulting in reduced ciliation. Consistent with impaired nucleation, altered structure and decreased growth speed of MTs during ciliogenesis, we also observed reduced migration of ciliated centrosomes to the plasma membrane, a movement driven by pushing forces from MTs nucleation, bundling and polymerization^32^. However, the impact of this molecular defect on ciliation and disease pathology is hard to evaluate. In principle, it could have no effect at all if the slowly migrating cilia eventually reach the cell surface during ciliogenesis *in vivo*. Conversely, it is also possible that cilia are selectively disassembled if they remain for too long in the cytosol where they are not fully functional. In this case, defective migration would also contribute to the observed reduction in ciliation. For clarity, the proposed model for the molecular mechanism by which TUBB G08S affects ciliogenesis is summarized in **Figure S7**.

While a similar molecular mechanism is conceivable for the E108K variant which also shows partial incorporation into cellular MTs, the E401K variant possibly acts through a different mechanism. TUBB E401K was previously shown to impair the chaperone-mediated assembly of αβ tubulin dimers and to be unable to incorporate into MTs^33^. These features suggested a loss-of-function mode of action for E401K, which was partially confirmed by phenotypic similarities in the brain of E401K mutant mice and heterozygous knock-out mice, both showing excessive apoptosis of cortical upper layer neurons leading to microcephaly^45^. However, some gain-of-function phenotypes of the E401K variant were also observed in the same studies. Overexpression of the mutant protein in developing mouse brain resulted in aberrant neuronal migration and positioning^33^, while the presence of ectopic progenitors and defects in mitotic spindle orientation were observed in the embryonic cortex of E401K mutant mice but not of heterozygous knock-out mice^45^. These findings led to the hypothesis that the E401K variant acts through a complex dual mechanism where loss-of-function effects are associated to microcephaly while gain-of-function effects are linked to defective neuronal positioning and caused by undetected residual incorporation of the mutant protein into MTs or by a deleterious effect on the tubulin folding pathway^33,45^. We propose that reduced ciliation is due to the gain-of-function mode of action of TUBB E401K and show that minimal (∼2% of cells) incorporation of TUBB E401K into cellular MTs can occur, suggesting that the mutant protein is not intrinsically incapable of incorporating into MTs. Interestingly, we observed a striking counterintuitive direct correlation between the diffuse appearance of each TUBB variant upon overexpression and its gain-of-function effect on ciliation. This finding suggests a possible alternative interpretation of the MT incorporation assay, especially for variants that show residual, albeit minimal, incorporation. In such cases, the assay may reflect how stably the mutant tubulin is incorporated into MTs rather than its intrinsic ability to incorporate. In this scenario, which is not mutually exclusive with poorly incorporated gain-of-function variants impairing the tubulin folding pathway, the proportion of cells showing diffuse tubulin would indicate the severity of the unstable incorporation phenotype and, in turn, of the destabilizing effect on MTs. Although the three variants may act through different molecular mechanisms, it is likely that they all ultimately affect the early stages of ciliogenesis as our data argue against the presence of TUBB in the axoneme of human primary cilia, at least in the cell types used in this study. Indeed, primary skin fibroblasts from the patient showed normal cilia length and morphology, and V5-tagged wild-type TUBB did not localize to the ciliary axoneme when overexpressed in hTERT-RPE1 cells. Even though an axonemal role for TUBB in the developing brain cannot in principle be excluded, our work suggests that TUBB is present in the MT subpopulation involved in PCV trafficking and centrosome migration, shedding some light on the largely unexplored field of the roles of distinct tubulin isotypes in ciliogenesis.

In our overexpression screen, some pathogenic TUBB variants did not show any ciliation defect. These include all the CSC-KT causing mutations (Q15K and Y222F), which do not cause brain malformations, but also three variants associated to brain abnormalities in ClinVar (T221I, M299V and V353I). No functional evidence of pathogenicity or description of the patient’s brain is available for T221I, while M299V and V353I have been validated to cause microcephaly with structural brain abnormalities^33^. Although gain-of-function phenotypes have been reported for M299V and V353I, which caused defects in neuronal migration (both variants) and positioning (M299V) upon overexpression in developing mouse brain, their molecular mechanism of action is significantly different from that of the ciliation-impairing variants G308S, E401K and E108K as they have no (V353I) or moderate (M299V) impact on the tubulin folding pathway and show complete, stable incorporation into cellular MTs^33^. Considering the role for defects in MT structure, nucleation and growth rate in impairing ciliogenesis and the proposed model linking those defects to unstable incorporation of the mutant tubulins, it is conceivable that these stably incorporating variants cause subtler effects on MT properties and dynamics that do not affect ciliation, or that affect it too mildly for the sensitivity of our assay. Furthermore, our screen is limited by the assumption of a gain-of-function mode of action for the tested variants, as loss-of-function mutations would not show any phenotype upon overexpression in wild-type cells. The ability of *TUBB* haploid cells to ciliate normally indeed suggests that heterozygous loss-of-function mutations in *TUBB* cannot affect ciliation. However, we observed that *TUBB* haploid cells partially compensate for the deleted allele at the transcriptional level. The mechanism used by cells to sense tubulin levels is currently unknown, nor is it known if this partial compensation is crucial to restore normal ciliation in *TUBB* haploid cells. Therefore, the possibility of a heterozygous loss-of-function missense mutation that escapes the sensing mechanism and impairs ciliation, although unlikely, cannot in principle be excluded.

We propose defective ciliogenesis to contribute to disease pathogenesis in at least a subset of *TUBB*-related tubulinopathy patients, alongside other cellular phenotypes associated with MT dysfunction such as the mitotic and spindle defects described for the E401K variant^45^. The heterogeneity in clinical manifestations and molecular phenotypes of the pathogenic *TUBB* variants studied so far suggests that each mutation has a specific mode of action, where the relative contribution of different pathogenic mechanisms may vary^33,34,45^. Importantly, some interindividual variability in disease manifestation is not surprising, as tubulinopathies and neurodevelopmental ciliopathies are two genetically heterogeneous groups of disorders characterized by a complex genotype-phenotype correlation. For both, different mutations in the same gene can result in specific phenotypes, and different phenotypes or disease severity can occur in individuals carrying the same variant, suggesting an important role for genetic modifier or environmental factors^4,13^.

Our data link for the first time tubulinopathies and neurodevelopmental ciliopathies, two groups of disorders affecting brain development. We showed that some gain-of-function patient mutations in the *TUBB* gene affect ciliogenesis and can cause ciliopathy-like brain phenotypes, revealing a novel disease mechanism for TUBB-related tubulinopathies and a novel ciliopathy-associated gene. Moreover, these findings can prompt future investigations on the link between tubulinopathies and primary cilia beyond the *TUBB* gene. Indeed, six tubulin genes have been associated to neurodevelopmental defects to date (*TUBA1A*, *TUBB*, *TUBB2A*, *TUBB2B*, *TUBB3*, *TUBG1*)^4^, but the contribution of distinct tubulin isotypes to cilia formation and homeostasis is still unknown. Furthermore, while tubulinopathies are considered incurable due to their developmental nature^54^, primary cilia are also present in differentiated neurons of the adult brain, where they are crucial to maintain neuronal viability and connectivity^14,55^. Therefore, the involvement of primary cilia defects in the pathogenesis of tubulinopathies reveals a new potential point of intervention to improve disease outcome after birth.

## METHODS

### Subject recruitment and clinical evaluation

This study was approved by the Research Ethic Board of The Hospital for Sick Children, Toronto, Ontario, Canada. Informed written consent to participate in research was obtained from the patient, the patient’s family members and control subjects. Clinical data were obtained by direct examination of participants and retrospective chart review.

### Whole genome sequencing

Genomic library preparation and whole genome sequencing (WGS) of the participants’ DNA were conducted at The Centre for Applied Genomics (TCAG) at The Hospital for Sick Children, Toronto, Ontario, Canada. 100 ng of DNA was used as input material for library preparation using the Illumina TruSeq PCR free Library Prep Kit following the manufacturer’s recommended protocol. Libraries were validated on a Bioanalyzer DNA High Sensitivity chip to check for size and absence of primer dimers, and quantified by qPCR using Kapa Library Quantification Illumina/ABI Prism Kit protocol (KAPA Biosystems). Each sample was sequenced on one lane of an Illumina HiSeq X platform following Illumina’s recommended protocol to generate paired-end reads of 150 bases in length. Each lane generated 112.5 Gb of data for ∼30X coverage of the human genome.

### Whole genome sequencing analysis and variant identification

WGS reads were aligned to the UCSC hg19 human reference genome assembly. Bioinformatic analysis of genetic variants was performed using a pipeline established in the Care4Rare project^56^. Namely, the germline variant calling pipeline used is implemented in the bcbio-nextgen framework (https://bcbio-nextgen.readthedocs.io/en/latest/index.html) and it is validated based on the Genome Analysis Toolkit’s (GATK) best practices^57^ and the standard NA12878 validation sample. GATK 4.0.7.0 was used, and variants in all the family members’ samples were called simultaneously. Variants were annotated using Ensembl Variant Effect Predictor (VEP)^58^, GEMINI^59^ and Vcfanno^60^. A detailed description of variants annotation can be found at https://docs.google.com/document/d/1zL4QoINtkUd15a0AK4WzxXoTWp2MRcuQ9l_P9-xSlS4/edit?usp=sharing.

To prioritize variants, we first restricted our analysis to gene coding regions and variants rare in the general population (gnomAD^61^ popmax < 1%). Among the resulting 1328 variants, we selected those that segregate with the disease phenotype in the family following both dominant and recessive modes of inheritance. No biallelic variant was found to segregate with the disease phenotype in the family under a recessive model. The dominant model with a *de novo* hypothesis, where a heterozygous variant is present in the proband and absent in parents and siblings, resulted in 12 variants. Of those, a missense variant in *TUBB* (NM_178014.3: c.922G>A/p.G308S) became our top candidate since only the *TUBB* gene had an OMIM (https://omim.org) description (#615771: cortical dysplasia, complex, with other brain malformations 6, autosomal dominant - #156610: symmetric circumferential skin creases, congenital 1, autosomal dominant). We also filtered all coding and rare (gnomAD^61^ popmax < 1%) non-coding variants in the 201 genes of the “Rare multisystem ciliopathy disorders” panel downloaded from https://panelapp.genomicsengland.co.uk/panels/150. This resulted in 21 coding variants, none of which segregated in the family according to the phenotype, and 348169 non-coding variants. Non-coding variants were filtered under a recessive model of inheritance (Hom in proband, Het in parents, non-Hom in siblings; OMIM gene description is present; OMIM inheritance mode is autosomal recessive). This filter resulted in 169 variants. We reviewed the variants manually, removing low coverage calling artifacts and, based on the variant annotations, we reached the conclusion that none of the variants was contributing to the disease phenotype.

Additionally, we generated a more stringent variant report containing ultra-rare (gnomAD^61^ popmax < 0.5%, frequency in the internal Care4Rare database < 5), high quality (variant quality score ≥ 1000, alt depth ≥ 20 in one of the samples) variants not restricted by a gene panel. This filter resulted in 40911 coding and non-coding variants, of which 3479 were in 1240 genes present in OMIM. We reviewed these variants using both dominant and recessive models and have not identified additional candidates that segregate in the family according to the phenotype.

### Antibodies

For immunostaining experiments, the following primary antibodies were used: mouse monoclonal anti-α-tubulin (DM1A, Sigma-Aldrich T6199, 1:500 dilution. Figure 2), mouse monoclonal anti-α-tubulin (DM1A, Sigma-Aldrich T9026, 1:10000 dilution. Figure 3 and 6), chicken polyclonal anti-V5 tag (Abcam ab9113, 1:500 dilution), mouse monoclonal anti-tyrosinated tubulin (TUB-1A2, Sigma-Aldrich T9028, 1:500 dilution), mouse monoclonal anti-γ-tubulin (GTU-88, Sigma-Aldrich T6557, 1:500 dilution for figure 3, 1:2000 dilution for figure 4, 5 and 7), rabbit polyclonal anti-ninein (Sigma-Aldrich ABN1720, 1:300 dilution), rabbit polyclonal anti-ARL13B (Proteintech 17711-1-AP, 1:300 dilution), mouse monoclonal anti-acetylated tubulin (6-11B-1, Sigma-Aldrich T7451, 1:500 dilution), rabbit polyclonal anti-CEP135 (Sigma-Aldrich SAB4503685, 1:500 dilution).

For immunoblotting, the following primary antibodies were used: mouse monoclonal anti-β-tubulin (AA2, Sigma-Aldrich T8328, 1:500 dilution), mouse monoclonal anti-β-actin (AC-15, Sigma-Aldrich A5441, 1:1000 dilution).

The following secondary antibodies were used: goat anti-rabbit IgG (H+L) Alexa Fluor™ 488 (ThermoFisher A-11008), goat anti-rabbit IgG (H+L) Alexa Fluor™ 555 (ThermoFisher A-21429), goat anti-mouse IgG (H+L) Alexa Fluor™ 488 (ThermoFisher A-11001), goat anti-mouse IgG (H+L) Alexa Fluor™ 555 (ThermoFisher A-21422), goat anti-chicken IgY (H+L) Alexa Fluor™ 488 (ThermoFisher A-11039), goat anti-mouse IgG (H+L) HRP-conjugated (Jackson ImmunoResearch 115-035-146).

### Plasmids

For gene editing experiments, the following plasmids were used: pSpCas9(BB)-2A-Puro (PX459) V2.0, which was a gift from Feng Zhang (Addgene plasmid no. 62988); BPK1520_blastR, the assembly of which was described previously^46^; pCMV-PE2, which was a gift from David Liu (Addgene plasmid # 132775); pU6-pegRNA-GG-acceptor, which was a gift from David Liu (Addgene plasmid # 132777). Plasmids for gRNA expression were generated by ligating annealed oligonucleotides into the linearized PX459 or BPK1520_blastR vectors. Plasmids for prime editing guide RNA (pegRNA) expression were generated by Golden Gate assembly of annealed oligonucleotides into the pU6-pegRNA-GG-acceptor vector, as previously described^37^.

For overexpression experiments, the following plasmids were used: pEGFP-N1 expressing human EB1-GFP (JB131), which was a gift from Tim Mitchison and Jennifer Tirnauer (Addgene plasmid # 39299); pCMV3-TUBB:V5, which was generated for this study.

The pCMV3-TUBB:V5 plasmid was generated using two rounds of site-directed mutagenesis to remove the N-terminal GFPSpark tag from the commercially available pCMV3-GFPSpark:TUBB plasmid (Sino Biological HG11626-ANG) and insert a C-terminal V5 tag (GKPIPNPLLGLDST) with a two amino acids linker (GS). Single nucleotide polymorphism (SNP) TUBB variants were introduced into pCMV3-TUBB:V5 via site-directed mutagenesis. To preserve the sequence of the pCMV3 vector from unwanted mutations during PCR amplification, the whole TUBB:V5 insert was moved to the pUC19NotI cloning vector at the KpnI/NotI restriction sites and moved back to pCMV3 after mutagenesis.

All site site-directed mutagenesis experiments were performed using the Q5® Site-Directed Mutagenesis Kit (New England Biolabs E0554) following the manufacturer’s recommended protocol. The mutagenic primers used were designed using the online NEBaseChanger tool (https://nebasechanger.neb.com) and are listed in **Table S1**. In all experiments, the complete open reading frame (ORF) was sequenced after mutagenesis to ensure that only the desired mutation was present.

### Cell culture and transfection

hTERT-RPE1 cells (ATCC CRL-4000) and p53-null hTERT-RPE1 cells (a gift from Daniel Durocher) were cultured in DMEM/F12 1:1 Mix (Wisent Bioproducts 319-085-CL) supplemented with 10% heat-inactivated FBS (Wisent Bioproducts 080-450) and 1% penicillin-streptomycin solution (Wisent Bioproducts 450-201-EL), and maintained at 37 °C in a humidified atmosphere (5% CO_2_). Primary skin fibroblasts were cultured in Alpha MEM (Wisent Bioproducts 310-010-CL) supplemented with 10% heat-inactivated FBS (Wisent Bioproducts 080-450) and 1% penicillin-streptomycin solution (Wisent Bioproducts 450-201-EL), maintained at 37 °C in a humidifie atmosphere (5% CO_2_) and used in experiments below passage 10. Where indicated, serum starvation was performed by washing cells twice with pre-warmed PBS (Wisent Bioproducts 311-010-CL) and culturing them in serum-free Opti-MEM™ (Thermo Fisher Scientific 31985-062) for the indicated time.

Transfections of hTERT-RPE1 cells and p53-null hTERT-RPE1 cells were carried out using Lipofectamine™ 3000 Transfection Reagent (Thermo Fisher Scientific L3000008) following the manufacturer’s recommended protocol. Unless otherwise indicated, cells were transfected at ∼80% confluence 24 hours after plating and medium was replaced 6 hours after transfection. Only for the generation of the *TUBB* haploid cell model, hTERT-RPE1 cells were transfected using the Neon™ Transfection System (Thermo Fisher Scientific MPK5000) with a pulse voltage of 1050 V, a pulse width of 30 ms and a pulse number of 2.

### Cell lines generation

Primary fibroblast cell lines were established from a skin biopsy of the patient and an age- and sex-matched control subject at The Centre for Applied Genomics (TCAG), Tissue Culture Facility - Biobanking Services at The Hospital for Sick Children, Toronto, Ontario, Canada according to their standard protocols. The patient-derived cell line was authenticated by Sanger sequencing for the *TUBB* NM_178014.3: c.922G>A/p.G308S variant **(Figure 4a)**.

The TUBB G308S heterozygous hTERT-RPE1 cell line (TUBB^G308S^) was generated by prime editing 3b (PE3b)^37^. Consistent with previous findings that functional p53 lowers the efficiency of Cas9-mediated gene editing even in absence of double-strand breaks^62–64^, p53-null hTERT-RPE1 cells (a gift from Daniel Durocher) were needed to achieve an editing efficiency detectable via Sanger sequencing in the bulk cellular population after antibiotic selection. Cells were plated at 2 x 10^5^ cells/well on 12-well plates (Corning 3513) and transfected the following day as described above. 845 ng of the pegRNA-expression plasmid (pU6-pegRNA-GG-acceptor), 280 ng of the nicking gRNA (ngRNA)- and blasticidin resistance-expression plasmid (BPK1520_blastR), and 375 ng of the prime editor-expression plasmid (pCMV-PE2) were cotransfected. At 24 hours post-transfection, blasticidin (Thermo Fisher Scientific A1113903) was added to the cells at 30 μg/ml in complete medium and selection medium was replaced daily for the following 6 days. Next, cells were resuspended in FACS buffer (2% FBS, 2.5 mM EDTA in PBS) at a concentration of 1 x 10^6^ cells/well, filtered through a 40 μm cell strainer (VWR CA21008-949) and single-cell sorted using a MoFlo XDP cell sorter (Beckman Coulter) into a 96-well plate (Corning 3599) containing complete medium. Clonal lines were expanded, and genomic DNA was extracted upon passaging for genotyping. Genotyping was performed by PCR amplifying the target region, and sequencing the targeted locus. Attempts to generate this cell line by homology-directed repair (HDR) and base editing (BE) were unsuccessful.

The *TUBB* haploid cell line was generated in wild-type hTERT-RPE1 cells (ATCC CRL-4000). WGS data from hTERT-RPE1 cells were accessed from the NCBI Short Read Archive (experiment ID: SRX858057). The target heterozygous polymorphism in *TUBB* intron 1 (NM_178014.4: c.57+235G>C) was manually identified using the Integrative Genomics Viewer^65^ and its presence was confirmed via Sanger sequencing of genomic DNA **(Figure 7a-i)**. The target polymorphism was chosen as it generates or destroys an SpCas9 PAM sequence, allowing for allele-specific editing. The final selection of both gRNAs used in this experiment (the allele-specific one targeting *TUBB* intron 1, and the one targeting the intergenic region downstream of *TUBB*) was based on minimizing off-target sites, as determined by the CHOPCHOP v.3 software^66^. hTERT-RPE1 cells are endogenously resistant to puromycin because of the method used for their immortalization, and the *TUBB* haploid cell line was established before the gRNA- and blasticidin resistance-expression plasmid (BPK1520_blastR) was assembled in our laboratory^46^. Therefore, EGFP cotransfection was used to allow FACS-based enrichment of transfected cells. Cells were transfected using the Neon™ Transfection System (Thermo Fisher Scientific MPK5000) as described above, and plated at 1 x 10^5^ cells/well on 24-well plates (Corning 3524). The two gRNAs were cloned into the gRNA- and SpCas9-expression vector PX459 and transfected in equal proportions (1000 ng each) into cells, together with a 7.5 kb CMV-EGFP expression plasmid assembled in our laboratory (200 ng). At 48 hours post-transfection, EGFP positive cells were sorted using a MoFlo XDP cell sorter (Beckman Coulter), replated and allowed to recover for 5 more days. At 1 week post-transfection, cells were single-cell sorted into a 96-well plate containing complete medium, as described above. Clonal lines were expanded, and genomic DNA was extracted upon passaging for genotyping. Genotyping was performed by amplifying and sequencing the predicted deletion junction formed by the joining of the two cut-sites **(Figure 7a-ii and 7a-iii)**. To confirm the successful haploidization of the target area and ensure that the deleted fragment was not retained elsewhere in the genome, the copy number of the targeted region was quantified by digital droplet PCR as described below **(Figure 7b)**.

The sequences of the pegRNAs, ngRNAs and gRNAs used in this study are listed in **Table S2.**

### Genomic DNA isolation and PCR amplification

Genomic DNA isolation was performed using the DNeasy Blood & Tissue Kit (Qiagen 69506) according to the manufacturer’s recommended protocol. PCR amplification was performed using CloneAmp™ HiFi PCR Premix (Takara Bio 639298) for genotyping the *TUBB* locus and using DreamTaq Green PCR Master Mix 2X (Thermo Fisher Scientific K1082) in all other instances, following the manufacturer’s recommended protocols.

The sequences of the primers used for PCR amplification are listed in **Table S3**.

### RNA isolation, semiquantitative and quantitative RT-PCR

Total RNA isolation was performed using the RNeasy Mini Kit (Qiagen 74106) according to the manufacturer’s recommended protocol. 1 μg of total RNA was reverse transcribed to cDNA using SuperScript™ III Reverse Transcriptase (Thermo Fisher Scientific 18080044) following the manufacturer’s recommended protocol (20 μl reaction volume).

For semiquantitative RT-PCR, PCR amplification was performed on a 1:5 dilution of the template cDNA for 25 cycles using DreamTaq Green PCR Master Mix 2X (Thermo Fisher Scientific K1082) following the manufacturer’s recommended protocol.

Quantitative RT-PCR (qPCR) was performed on a 1:100 dilution of the template cDNA using PowerUp™ SYBR™ Green Master Mix (Thermo Fisher Scientific A25776) in a QuantStudio™ 3 Real-Time PCR System (Thermo Fisher Scientific, Applied Biosystems A28136). The relative expression levels of *TUBB* were compared using the ΔΔCt method, with *GAPDH* used as housekeeping control gene for normalization. The primers for *TUBB* are located within the deleted region in the *TUBB* haploid cell line, were previously used in qPCR^67^, and their amplification efficiency was verified by generating a standard curve with four 1:10 serial dilutions of the template cDNA.

The sequences of the primers used for semiquantitative and quantitative RT-PCR experiments are listed in **Table S3**. To prevent amplification of genomic DNA, at least one primer of each pair was designed to span an exon-exon boundary in the target mRNA.

### Protein isolation and immunoblotting

To immunoblot for total β-tubulin and β-actin, TUBB^WT^ and TUBB^G308S^ cells were plated on 6-well plates (Corning 3506) and grown overnight in complete medium. Cells were resuspended in ice-cold 1X RIPA Lysis Buffer IV with Triton-X-100 pH 7.4 (Bio Basic RB4478) supplemented with Protease Inhibitor Cocktail (Sigma-Aldrich P8340). Protein concentration was determined using the Pierce™ BCA Protein Assay Kit (Thermo Fisher Scientific 23225) following the manufacturer’s recommended protocol. Next, an equal volume of 2X Laemmli Sample Buffer (Bio-Rad 1610737) was added to the samples and lysates were boiled for 10 minutes. Equal amounts of protein lysates were separated on 4-20% Mini-PROTEAN® TGX™ Precast Protein Gels (Bio-Rad 4561096) in 1X Tris/Glycine/SDS buffer (Bio-Rad 1610732) and electro-blotted to nitrocellulose membranes in 1X Tris/Glycine buffer (Bio-Rad 1610734) supplemented with 20% methanol (Fisher Scientific A412-4). Membranes were blocked in 1X TBST [Tris-Buffered Saline (50 mM Tris pH 7.5, 150 mM NaCl) with 0.1% Tween® 20 detergent (Fisher Scientific BP337-500)] containing 5% non-fat powdered milk (Bio Basic NB0669) for 1 hour at room temperature. Membranes were probed at 4 °C overnight with the primary antibodies indicated above, diluted in blocking buffer. Next, membranes were washed in 1X TBST and incubated for 1 hour at room temperature with the HRP-conjugated anti-mouse secondary antibody indicated above (1:10000 dilution in blocking buffer). Chemiluminescence signals were detected using SuperSignal™ West Pico PLUS Chemiluminescent Substrate (Thermo Fisher Scientific 34580) and acquired using a ChemiDoc Imaging System (Bio-Rad). Immunoblots were exposed for durations ranging from 0.3 to 2 seconds without saturating the camera’s pixels.

### Copy number analysis by digital droplet PCR

Genomic copy number estimation of *TUBB* was conducted at The Centre for Applied Genomics (TCAG) at The Hospital for Sick Children, Toronto, Ontario, Canada. Copy number analysis was performed on the QX200 Droplet Digital PCR System (Bio-Rad) using a Custom TaqMan™ Copy Number Assay (Thermo Fisher Scientific 4400294). The primer sequences are shown in **Table S4**. 50 ng of genomic DNA was digested with 5U of DraI (New England Biolabs R0129) in a 3 μl reaction with a 1-hour incubation at 37 °C and no enzyme denaturation. The 20 μl copy number reaction mix consisted of 10 μl of 2X ddPCR™ Supermix for Probes (Bio-Rad 1863026), 1 μl of the copy number target assay (labeled with FAM), 1 μl of the VIC-labelled copy number reference assay (*RNAseP*, Thermo Fisher Scientific 4403326 or *NPC1*, **Table S4**), 5 μl of water and 3 μl of 16.7 ng/μl digested genomic DNA. Cycling conditions for the reaction were 95 °C for 10 min, followed by 45 cycles of 94 °C for 30 seconds and 60 °C for 1 minute, 98 °C for 10 minutes and finally a 4 °C hold on a Life Technologies Veriti thermal cycler. Data were analyzed using QuantaSoft v1.4 (Bio-Rad).

### Immunofluorescence

For indirect immunofluorescence, cells were plated on glass coverslips (Electron Microscopy Sciences 72230-01) in 12-well plates (Corning 3513) and treated according to each experiment. Cells were then washed once with PBS (Wisent Bioproducts 311-010-CL) and fixed in methanol (Fisher Scientific A412-4) at -20 °C for the time indicated for each experiment. Next, coverslips were blocked with 5% FBS (Figure 2, 4, 5, 7, 8 and S5) or 3% BSA, 1% FBS (Figure 3, 6 and S4) in PBST [PBS with 0.05% Tween® 20 detergent (Fisher Scientific BP337-500)]. Coverslips were subsequently incubated with the primary antibodies indicated above diluted in blocking buffer, in a humidified chamber overnight at 4 °C. Coverslips were then washed three times in PBST and incubated in a humidified chamber with the appropriate Alexa Fluor™-conjugated secondary antibody (1:1000 dilution in blocking buffer) for 1 hour at room temperature, and counterstained for 5 minutes with 1 μg/ml DAPI (Thermo Fisher Scientific 62248) or Hoechst 34580 (Thermo Fisher Scientific H21486) in PBS. Following three washes in PBST, coverslips were mounted on microscope glass slides (VWR 48312-401) using ProLong Gold Antifade Mountant (Thermo Fisher Scientific P36930).

### Fluorescent imaging

Whole slide scans were acquired on a Pannoramic 250 Flash III slide scanner (3DHistech) using a 40X 0.95 NA air objective (Zeiss). The instrument was operated in extended focus mode (seven focal planes spanning 5 μm axial distance) to capture the entire cell volume and all primary cilia that may lie on different focal planes.

Epifluorescence images were acquired on a Zeiss Axio Observer Z1 inverted microscope using a Axiocam 506 mono camera (Zeiss) with a 40X 1.3 NA oil immersion objective (Zeiss), configured using Zeiss Zen 3.1 image acquisition software.

Confocal images in figure 2, 4, 5, 7, 8 and S5 were acquired on a Leica SP8 Lightning confocal microscope (Leica Microsystems) using HyD detectors in combination with a 63X 1.3 NA oil immersion objective (Leica Microsystems) controlled by Leica Application Suite X (LAS X) image acquisition software. Confocal images in figure 3, 6 and S4 were acquired on a Quorum WaveFX-X1 spinning disk confocal microscope (Quorum Technologies) using a cooled electron-multiplying charged-coupled device (EM-CCD) camera (Hamamatsu) with a 40X 1.3 NA or a 63X 1.4 NA oil immersion objective, configured using MetaMorph® image acquisition software (Molecular Devices). Unless otherwise indicated, consecutive Z-stacks were acquired at Nyquist intervals to capture the entire cell volume and all primary cilia that may lie on different focal planes. Unless otherwise indicated, maximum intensity projections of all the Z-planes are shown in the figures. All live-cell imaging was acquired on a Quorum WaveFX-X1 spinning disk confocal microscope with a 63X 1.4 NA oil immersion objective. Cells were kept at 37 °C with 5% CO_2_ throughout imaging.

The Pannoramic 250 Flash III slide scanner and the Leica SP8 Lightning microscope are housed in The Imaging Facility at The Hospital for Sick Children, Toronto, Ontario, Canada. The Quorum WaveFX-X1 microscope is housed in the Centre for the Neurobiology of Stress, University of Toronto Scarborough, Ontario, Canada.

### Fluorescence intensity measurements

Measurements of fluorescence intensity of MTs, EB1-GFP comets, ninein, and γ−tubulin were generated by using the selection tools in Fiji^68^ to encompass intensities from single frames or maximum intensity projections, followed by background intensity subtraction from each measurement.

### MT incorporation assay

MT incorporation assay was performed in wild-type hTERT-RPE1 cells (ATCC CRL-4000). Cells were plated at 2 x 10^5^ cells/well on glass coverslips in 12-well plates and transfected the following day with either wild-type or TUBB G308S pCMV3-TUBB:V5 plasmid, as described above. At 24 hours post-transfection cells were fixed in methanol at -20 °C for 5 minutes and processed for immunofluorescence as described above, without staining cell nuclei.

MT incorporation was assessed on whole slide scans visualized using CaseViewer 2.4 (3DHistech). For each coverslip, three different fields (ROI: regions of interest) were selected for analysis. To avoid sampling biases, the three ROIs were randomly selected across the whole coverslip and all the low and medium TUBB-V5 expressing cells in each ROI were scored. Analysis was performed at the same zoom level (90X) for all the samples and blind to sample identity. MT incorporation was visually quantified by assigning each transfected cell to the “Incorporated”, “Partially incorporated” or “Diffuse” category based on the filamentous, partially diffuse or diffuse appearance of the fluorescent signal in the V5 channel.

Representative images shown in figure 2 are maximum intensity projections of confocal Z-stacks encompassing the entire cell volume.

### MT dynamics assay

For EB1-GFP live imaging in complete medium, 3 x 10^5^ cells were plated into 35-mm glass-bottom single well dishes (MatTek P35G-1.5-14-C) and transiently transfected the following day with the EB1-GFP expressing plasmid, as described above. For EB1-GFP live imaging in serum-starved conditions, 4 x 10^5^ cells were plated into 35 mm glass-bottom single well dishes and serum starved the following day in Opti-MEM™ as described above, upon transient transfection with the EB1-GFP expressing plasmid. In both experiments cells were imaged the day after plasmid transfection, around and no later than 24 hours post-transfection.

The speed of EB1 comets was quantified from the maximum intensity projection of confocal time-lapse images of low and medium EB1-GFP expressing cells using the TrackMate plugin^69^ in Fiji^68^. Images were subjected to bleach correction using simple ratio method prior to track analysis utilizing LoG Detector and Simple LAP Tracker. The integrity of EB1-GFP tracks was verified individually. The number of EB1-GFP comets was quantified using the TrackMate^69^ plugin in Fiji^68^ from the maximum intensity projection of confocal Z-stack images of medium and low EB1-GFP expressing cells. To reduce biases from variations in cell shape, a fixed-sized area at the centrosomes was used to count the EB1-GFP comets. EB1-GFP comets were estimated as 1 µm-diameter spots that were further analyzed using LoG Detector.

### MT regrowth assay

MT regrowth assay was performed by depolymerizing the MT network by treating cells with 10 μM nocodazole (Sigma-Aldrich SML1665) in complete medium for 20 minutes at 37 °C with 5% CO_2_. After depolymerization, cells were washed extensively with PBS to remove the nocodazole and incubated with pre-warmed complete culture medium to induce MT nucleation and regrowth at 37 °C with 5% CO_2_. After 2 or 4 minutes of MT regrowth, cells were fixed in methanol at -20 °C for 10 minutes.

### MT curvature analysis

MT curvature analysis was generated using the Kappa plugin^70^ in Fiji^68^. Semiautomated tracking of individual MTs was done on confocal slices using open B-spline configurations. The initialization curve is fitted using point distance minimization algorithm.

### Ciliation assays

For primary skin fibroblasts, 2 x 10^5^ cells/well were plated on glass coverslips in 12-well plates. The following day, cells (∼95% confluence) were serum starved in Opti-MEM™ as described above for 72 hours to induce ciliation, before fixation in methanol at -20 °C for 20 minutes and processing for immunofluorescence.

For hTERT-RPE1 cells, 3 x 10^5^ cells/well were plated on glass coverslips in 12-well plates. The following day, cells (∼90% confluence) were serum starved in Opti-MEM™ as described above for 2, 4, 6 **(Figure 6a)**, 24 **(Figure 6b and S4)** or 48 **(Figure 5 and 7)** hours to induce ciliation, before fixation in methanol at -20 °C for 20 minutes and processing for immunofluorescence.

For the overexpression screen, wild-type hTERT-RPE1 cells (ATCC CRL-4000) were plated at 2 x 10^5^ cells/well on glass coverslips in 12-well plates. The following day, cells were transfected as described above with either wild-type or mutant pCMV3-TUBB:V5 plasmid. At 24 hours post-transfection (∼90% confluence) ciliation was induced by serum starving cells in Opti-MEM™ as described above for 24 hours, before fixation in methanol at -20 °C for 20 minutes and processing for immunofluorescence.

The percentage of ciliated cells, the length of primary cilia and MT incorporation in the overexpression screen were assessed on whole slide scans visualized using CaseViewer 2.4 or SlideViewer 2.5 (3DHistech). For each coverslip, three different fields (ROI: regions of interest) were selected for analysis. To avoid sampling biases, the three ROIs were randomly selected across the whole coverslip using only the DAPI or Hoechst channel, and all the cells in each ROI were scored. All analyses were performed at the same zoom level (90X) and blind to sample identity. The percentage of ciliated cells was calculated as the number of primary cilia divided by the total number of DAPI- or Hoechst-labeled cell nuclei, as scored manually using the Marker Counter tool in CaseViewer. The length of primary cilia was measured using the Measurement Annotation tool in CaseViewer, approximating primary cilia as linear structures. In the overexpression screen, only low and medium TUBB-V5 expressing cells were scored for the analysis of the transfected cell population. MT incorporation was visually quantified by assigning each transfected cell to the “Incorporated”, “Partially incorporated” or “Diffuse” category based on the filamentous, partially diffuse or diffuse appearance of the fluorescent signal in the V5 channel.

Representative images shown in the figures are maximum intensity projections of confocal Z-stacks encompassing the entire cell volume.

### Centrosome migration measurement

The migration distance of ciliated centrosomes was measured from confocal Z-stack slices (0.2 µm apart) encompassing the entire thickness of the cell. Stacks were displayed as XY and XZ views using XYZ projection tool in Fiji^68^. Straight line function was used on XZ projection to measure the distance between CEP135 and the apex of DAPI-stained nucleus.

### Protein structural analyses

All 3D protein structures were rendered using the PyMOL Molecular Graphics System version 2.0 (Schrödinger, LLC). Mutant protein stability was calculated as change in Gibbs folding free energy (ΔΔG) using FoldX software^71^ with default settings. ΔΔG was calculated for every possible glycine to serine substitution over the four indicated protein structures (PDB IDs: 4i4t, 5nqu, 5yl2, 5ca1).

### Cell cycle analysis

Cell cycle analysis of cycling and serum-starved TUBB^WT^ and TUBB^G308S^ hTERT-RPE1 cells was performed by flow cytometry using propidium iodide (PI) to measure DNA content. For cycling samples, 1.5 x 10^5^ cells/well were plated in complete medium in 12-well plates. For serum-starved samples, the same conditions as the ciliation assay were used: 3 x 10^5^ cells/well were plated in complete medium in 12-well plates, and serum starved in Opti-MEM™ for 48 hours the following day, as described above. In preparation for flow cytometry ∼2 x 10^6^ cells/sample were collected, resuspended in 50 ml eBioscience™ Flow Cytometry Staining Buffer (Thermo Fisher Scientific 00-4222-26) and fixed with 1 ml of ice-cold 80% ethanol overnight at 4°C. Fixed cells were then collected, incubated with 500 μl of 2 mg/ml RNase A (Qiagen 19101) in HBSS with calcium and magnesium (Thermo Fisher Scientific 14025-092) for 5 minutes at room temperature, and stained for 30 minutes at room temperature with 500 μl of 0.1 mg/ml PI (Thermo Fisher Scientific P3566) in HBSS 0.6% NP-40 (Sigma-Aldrich I8896). Cells were next filtered through a 40 μm cell strainer (VWR CA21008-949) and flow cytometry was performed using a BD LSR II analyzer (BD Biosciences). Analysis of PI was performed using 561 nm (yellow-green) laser excitation and 575/26 nm bandpass filter. Flow cytometry data were gated for single nuclei, analyzed, and plotted using FlowJo™ software version 10.8 (BD Biosciences).

### Statistical analysis

Data normality was assessed using the Anderson-Darling or D’Agostino-Pearson omnibus tests. For normally distributed data, statistical significance was determined using two-tailed Student’s t-test. For non-normal distributed data, a non-parametric Mann U Whitney test was used. All statistical analyses were conducted using GraphPad Prism version 9.4.1 (GraphPad Software) with p-value (p) < 0.05 considered statistically significant.

## Data Availability

All data produced in the present study are available upon reasonable request to the authors

## ACKNOWLEDGMENTS

We thank Paul Paroutis (The Imaging Facility, The Hospital for Sick Children, Toronto) for his assistance with whole slide scanning. We thank Emily Reddy (Flow Cytometry Facility, The Hospital for Sick Children, Toronto) for her help with flow cytometry data acquisition and analysis. We thank Daniel Durocher (Department of Molecular Genetics, University of Toronto) for sharing the p53-null hTERT-RPE1 cells. We thank Elise Héon (Genetics and Genome Biology Program, The Hospital for Sick Children, Toronto) for sharing the control primary skin fibroblasts and for helpful discussions. We thank Dan Doherty (Center for Integrative Brain Research, Seattle Children’s Research Institute) and Julie Van De Weghe (Department of Pediatrics, University of Washington) for extensive and helpful discussions throughout this project. We are grateful to all the current and past members of the Cohn laboratory and the Ivakine laboratory for helpful discussions and input.

This study was supported by the Nicol Family Foundation, the SickKids Foundation, and by a grant from the Rare Disease Foundation and the BC Children’s Hospital Foundation (grant #2808 to A.M.).

A.M. is a recipient of an Ontario Trillium Scholarship (OTS). S.O. is a recipient of a University of Toronto Provost’s Postdoctoral Award. Research in R.E.H. laboratory is supported by a grant from the Canadian Institutes of Health Research (CIHR, grant #PJT-166084). The Centre for the Neurobiology of Stress at University of Toronto Scarborough is supported by a grant from the Canada Foundation for Innovation (CFI, grant #493864).

