## Supplementary Figures for "Brain development mutations in the β-tubulin TUBB result in defective ciliogenesis"

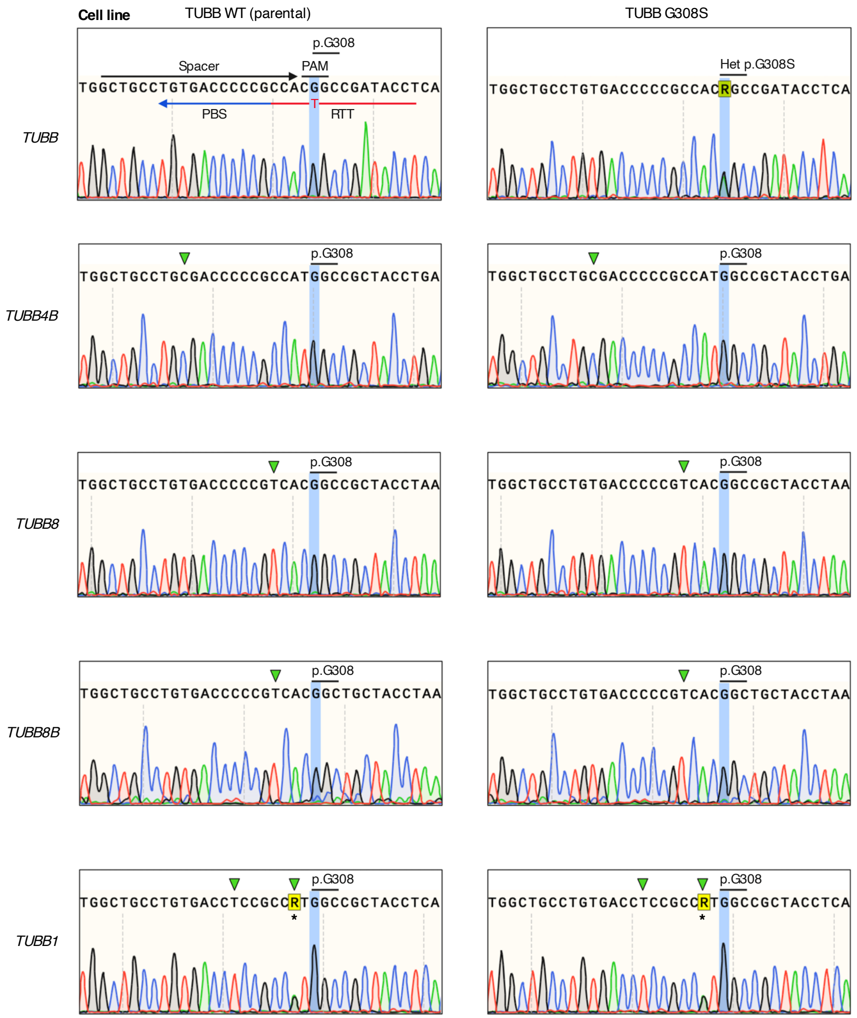


* Known heterozygous single nucleotide polymorphism in hTERT RPE-1 (ATCC CRL-4000) cells [NCBI Short Read Archive (SRA)-Experiment ID: SRX858057]

**Figure S1: Heterozygous TUBB$^{G308S}$ hTERT-RPE1 cells are not edited at the most likely off-target sites.** Sanger sequencing electropherograms of the target site (*TUBB* locus; upper panel) and the four most likely predicted protein-coding off-target sites (*TUBB4B, TUBB8, TUBB8B, TUBB1* loci; lower panels) in the generated heterozygous TUBB$^{G308S}$ cell model and parental TUBB$^{WT}$ cells. The target base in the *TUBB* locus and the corresponding base in the off-target loci are highlighted in blue. The analyzed off-target sites correspond to protein-coding regions of the human genome showing single or double mismatches with the spacer sequence of the used prime editing guide RNA (pegRNA). Green triangles indicate mismatch sites.

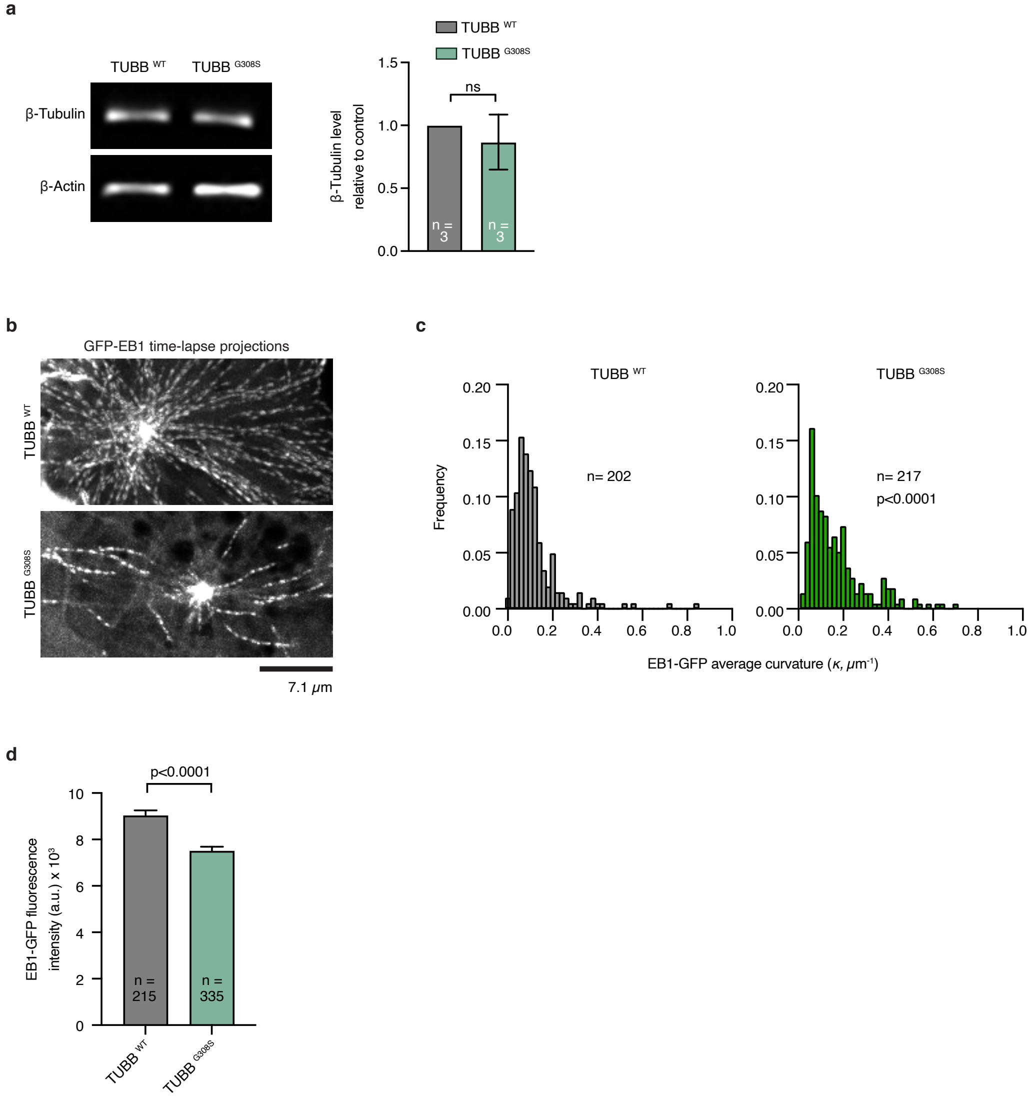

a
TUBB WT
TUBB G308S
β-Tubulin
β-Actin
β-Tubulin level relative to control
1.5
1.0
0.5
0.0
ns
n = 3
n = 3
b
GFP-EB1 time-lapse projections
TUBB WT
TUBB G308S
7.1 μm
c
TUBB WT
TUBB G308S
Frequency
0.20
0.15
0.10
0.05
0.00
n= 202
n= 217
p<0.0001
0.0 0.2 0.4 0.6 0.8 1.0
EB1-GFP average curvature (κ, μm⁻¹)
d
p<0.0001
EB1-GFP fluorescence intensity (a.u.) x 10³
10
8
6
4
2
0
n = 215
n = 335
TUBB WT
TUBB G308S

**Figure S2: Heterozygous TUBB$^{G308S}$ hTERT-RPE1 cells nucleate fewer microtubules that show excessive curvature.** (a) Representative immunoblot of total cell lysates from TUBB$^{WT}$ and TUBB$^{G308S}$ hTERT-RPE1 cells probed for β-tubulin and β-actin. Bar graph shows β-tubulin signal intensity normalized to β-actin and plotted relatively to TUBB$^{WT}$ cells. Data from three experiments are represented as mean ± standard deviation. P-value was calculated using an unpaired t-test. ns = not statistically significant ($p > 0.05$). (b) Representative spinning disk confocal images of TUBB$^{WT}$ and TUBB$^{G308S}$ cells expressing EB1-GFP. EB1-GFP projections were generated from maximum intensity projection of 12 frames captured with 4 seconds intervals. Growing microtubule ends appear as bright tracks. (c) Histogram of the frequency of the average curvature of EB1-GFP projections in TUBB$^{WT}$ and TUBB$^{G308S}$ cells. P-value was calculated using Mann U Whitney test. (d) Bar graph of the mean fluorescence intensity of EB1-GFP comets in transiently transfected TUBB$^{WT}$ and TUBB$^{G308S}$ cells. Data are represented as mean ± standard error of the mean. P-value calculated using an unpaired t-test.

**a**

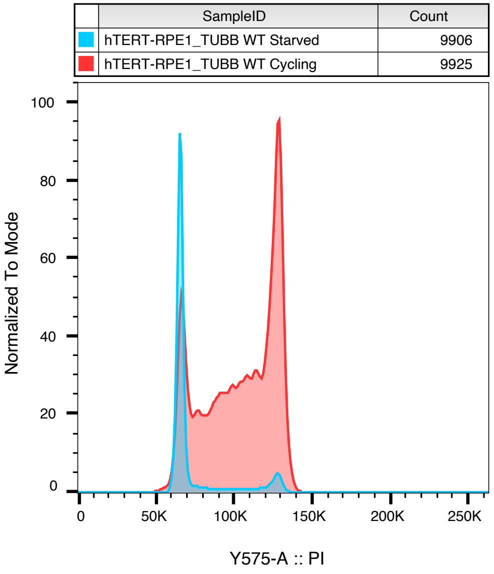
SampleID
Count
hTERT-RPE1_TUBB WT Starved
9906
hTERT-RPE1_TUBB WT Cycling
9925
Normalized To Mode
0
20
40
60
80
100
0
50K
100K
150K
200K
250K
Y575-A :: PI

**b**

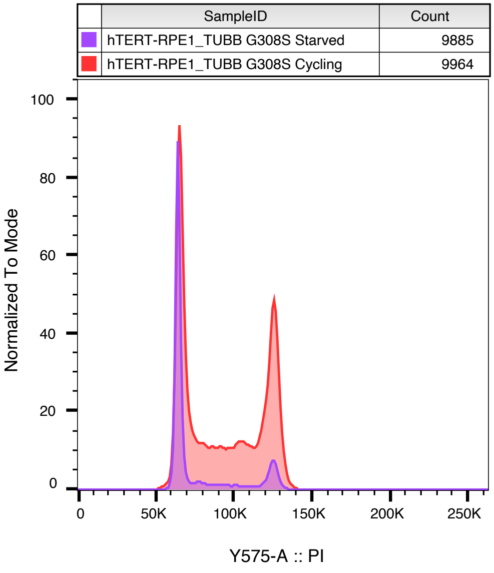
SampleID
Count
hTERT-RPE1_TUBB G308S Starved
9885
hTERT-RPE1_TUBB G308S Cycling
9964
Normalized To Mode
0
20
40
60
80
100
0
50K
100K
150K
200K
250K
Y575-A :: PI

**c**

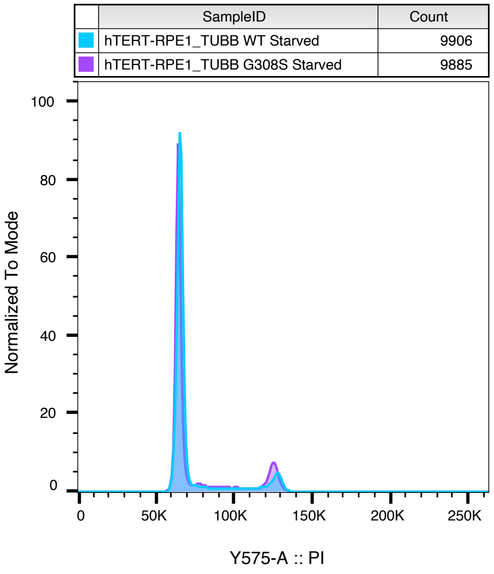
SampleID
Count
hTERT-RPE1_TUBB WT Starved
9906
hTERT-RPE1_TUBB G308S Starved
9885
Normalized To Mode
0
20
40
60
80
100
0
50K
100K
150K
200K
250K
Y575-A :: PI

**Figure S3: Heterozygous TUBB$^{G308S}$ cells exit the cell cycle as effectively as TUBB$^{WT}$ cells upon serum withdrawal.** Cell cycle profiles of TUBB$^{WT}$ and TUBB$^{G308S}$ hTERT-RPE1 cells, cycling or after 48 h serum withdrawal to induce quiescence, obtained by flow cytometry analysis using propidium iodide (PI) as a stoichiometric DNA binding dye to measure DNA content. Data are represented as histograms of PI fluorescence intensity, normalized to mode to account for differences in cell number across the samples. (a) Overlaid histograms of cycling (*red*) and serum-starved (*blue*) TUBB$^{WT}$ cells. (b) Overlaid histograms of cycling (*red*) and serum-starved (*purple*) TUBB$^{G308S}$ cells. (c) Overlaid histograms of serum-starved TUBB$^{WT}$ cells (*blue*) and serum-starved TUBB$^{G308S}$ cells (*purple*).

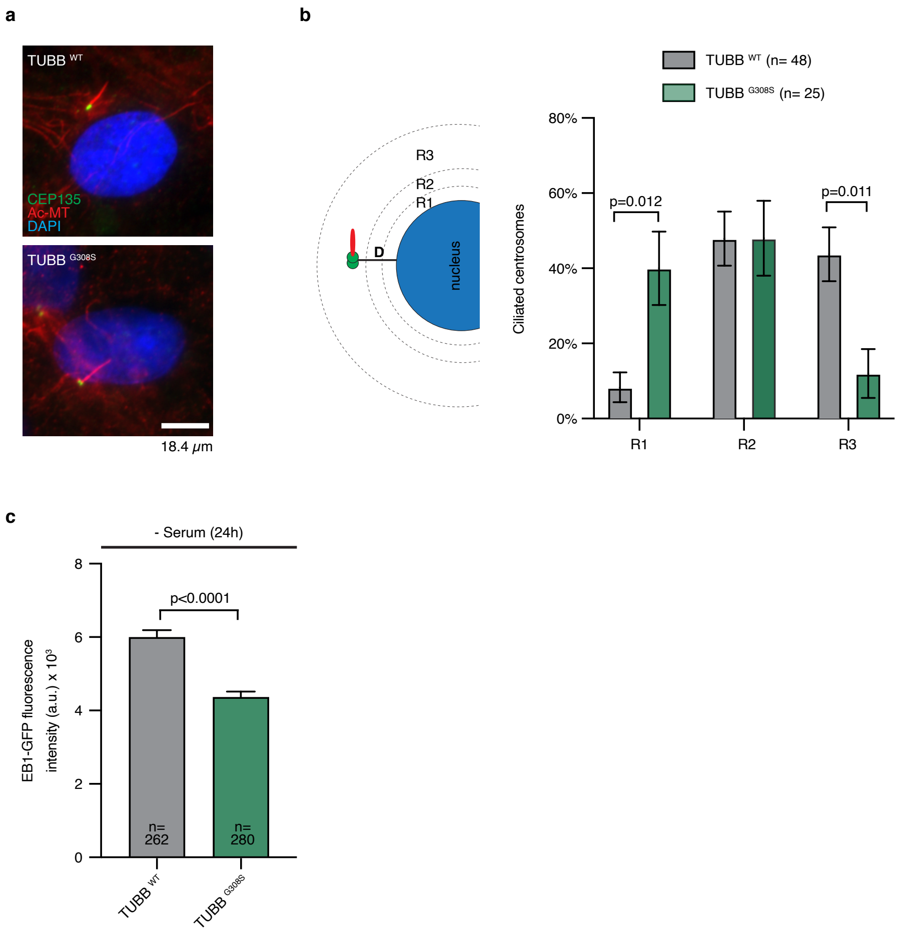
a
TUBB WT
CEP135
Ac-MT
DAPI
TUBB G308S
18.4 μm
b
R3
R2
R1
D
nucleus
TUBB WT (n= 48)
TUBB G308S (n= 25)
Ciliated centrosomes
80%
60%
40%
20%
0%
p=0.012
p=0.011
R1
R2
R3
c
- Serum (24h)
EB1-GFP fluorescence intensity (a.u.) x 10³
8
6
4
2
0
p<0.0001
n= 262
n= 280
TUBB WT
TUBB G308S

**Figure S4: Heterozygous TUBB$^{G308S}$ hTERT-RPE1 cells show defective primary cilia migration.** (a) Representative images of ciliated TUBB$^{WT}$ and TUBB$^{G308S}$ cells fixed after 24 h serum withdrawal and immunostained for the centrosomal marker CEP135 (*green*), acetylated-$\alpha$-tubulin (*red*) and DNA (*blue*) to label the nucleus. (b) Schematic representation showing the area around the nucleus of a ciliated cell. Distance (D) was measured between ciliated CEP135-positive centrosomes and the nearest margin of the nucleus. The following radii (R) were considered, from the nearest margin of the nucleus: R1 < 1 µm; 1 µm < R2 < 2 µm; R3 > 2 µm. Bar graph of the percentage of ciliated centrosomes observed within each designated region in TUBB$^{WT}$ and TUBB$^{G308S}$ cells after 24 h serum withdrawal. Data are represented as mean $\pm$ standard error of the mean. P-value was calculated using two-way ANOVA followed by Sidak's multiple comparisons test. Non-statistically significant ($p > 0.05$) comparisons are not shown. (c) Bar graph of the mean fluorescence intensity of EB1-GFP comets in transiently transfected TUBB$^{WT}$ and TUBB$^{G308S}$ cells after 24 h serum withdrawal. Data are represented as mean $\pm$ standard error of the mean. P-value was calculated using Mann U Whitney test.

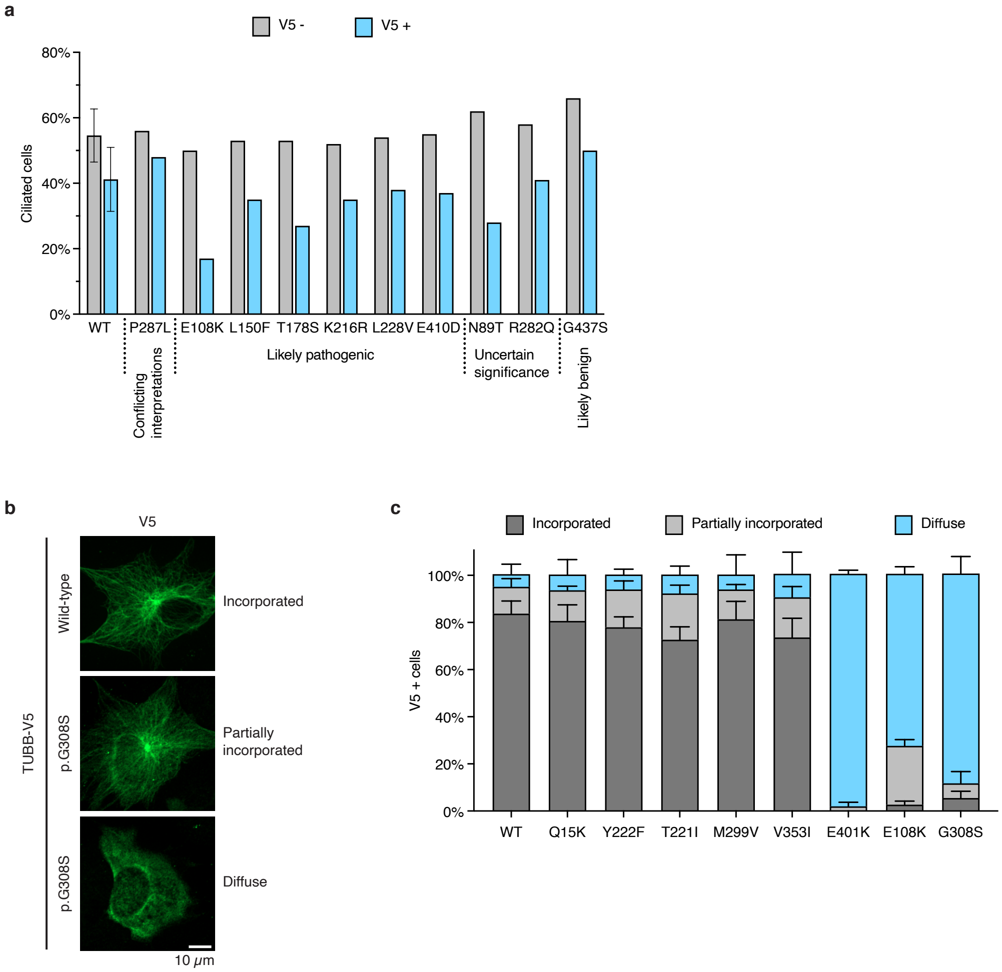
a
V5 -
V5 +
Ciliated cells
80%
60%
40%
20%
0%
WT
P287L
E108K
L150F
T178S
K216R
L228V
E410D
N89T
R282Q
G437S
Conflicting interpretations
Likely pathogenic
Uncertain significance
Likely benign
b
V5
TUBB-V5
Wild-type
p.G308S
p.G308S
Incorporated
Partially incorporated
Diffuse
10 μm
c
Incorporated
Partially incorporated
Diffuse
V5 + cells
100%
80%
60%
40%
20%
0%
WT
Q15K
Y222F
T221I
M299V
V353I
E401K
E108K
G308S

**Figure S5: A subset of clinically relevant dominant mutations in TUBB affects ciliation and shows reduced incorporation into microtubules in hTERT-RPE1 cells.** (a) Ciliation frequency of cells overexpressing wild-type or mutant (P287L, E108K, L150F, T178S, K216R, L228V, E410D, N89T, R282Q, G437S) TUBB-V5 after 24 h serum withdrawal. Percentage of cells showing ARL13B-positive cilium was measured in the transfected (V5-positive) and non-transfected (V5-negative) cell populations ($n \geq 200$ cells for V5-positive population, $n = 400$ cells for V5-negative population). For each mutation, clinical significance from the ClinVar database is shown. The graph presents pooled data from single-replicate preliminary experiments on TUBB variants with undetermined clinical significance at the time of this study. For the wild-type control sample, data from five experiments are represented as mean ± standard deviation. (b) Representative images of hTERT-RPE1 cells overexpressing wild-type or mutant (G308S) TUBB-V5 from the experiment in Figure 8. Cells were fixed after 24 h serum withdrawal and immunostained for V5 (*green*) and the ciliary marker ARL13B (*not shown*). Examples of complete microtubule incorporation, partial microtubule incorporation, and diffuse distribution of TUBB-V5 are shown. (c) Percentage of V5-positive cells from the experiment in Figure 8 showing complete microtubule incorporation, partial microtubule incorporation, and diffuse distribution of TUBB-V5 as determined by the appearance of the V5-signal. Pooled data from at least three experiments ($n \geq 200$ cells) are represented as mean ± standard deviation.

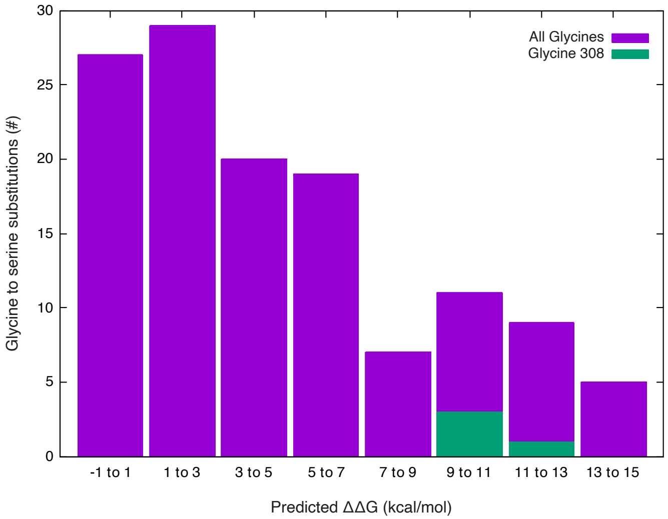

All Glycines
Glycine 308
Glycine to serine substitutions (#)
0
5
10
15
20
25
30
-1 to 1
1 to 3
3 to 5
5 to 7
7 to 9
9 to 11
11 to 13
13 to 15
Predicted ΔΔG (kcal/mol)

**Figure S6: The G308S mutation is predicted to destabilize TUBB fold.** *In silico* protein stability analysis on 4 beta-tubulin structures from 3 species (PDB IDs: 4i4t, 5nqu, 5yl2, 5ca1). The effect of every possible glycine to serine substitution was evaluated (software: FoldX). Combined results are shown.

$\Delta\Delta G = \Delta G\ (\text{folding})_{\text{mutant}} - \Delta G\ (\text{folding})_{\text{wild-type}}$ ; $\Delta\Delta G > 0$ if the mutation is destabilizing

Serum-starved conditions

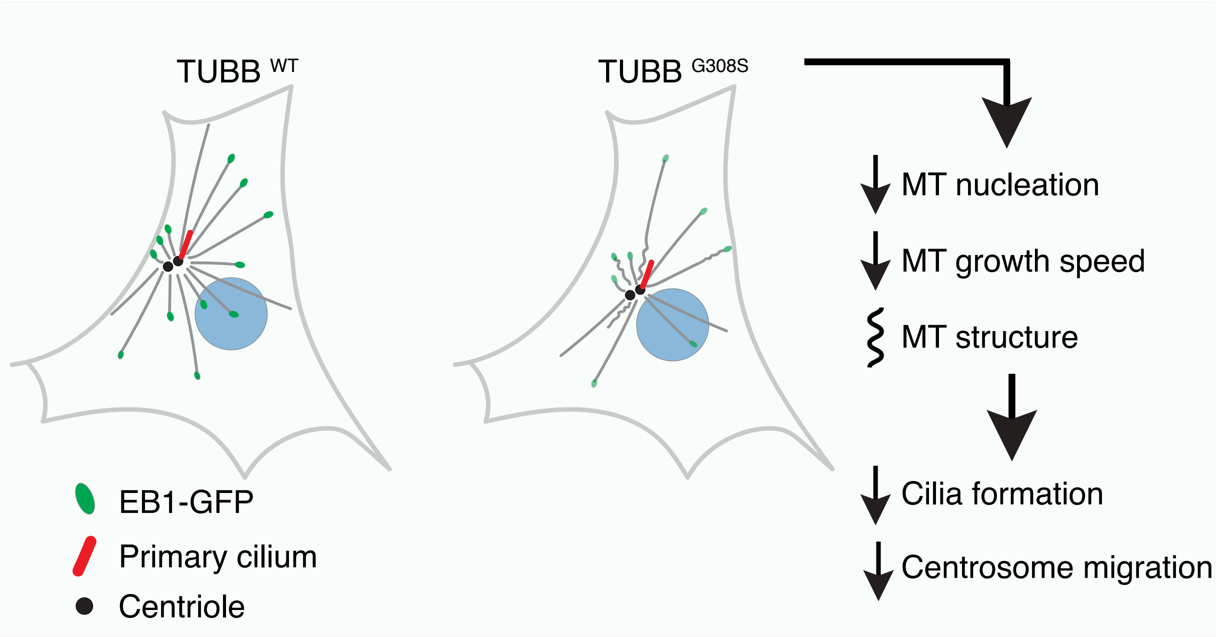

**Figure S7: Possible model for the effects of TUBB G308S on ciliogenesis.** In serum-starved $\text{TUBB}^{\text{G308S}}$ hTERT-RPE1 cells, the reduced nucleation, altered (wavy and buckled) structure, and decreased growth speed of MTs (*grey*) lead to impaired primary cilia formation and centrosome migration during ciliogenesis.

Primary cilia formation depends on the dynein-mediated transport of pre-ciliary vesicles (PCVs) to the mother centriole in the early stages of ciliogenesis[28,30]. The reduced MT density and the structural abnormalities of the existing MTs likely affect PCVs transport, as variations in MT structure such as bending and buckling have been shown to contribute to motor-mediated MT transport and influence vesicle motion[53]. Moreover, dynein recruitment to MT plus-end is likely impaired in $\text{TUBB}^{\text{G308S}}$ cells. It depends on the previous recruitment of EB1 (*green*)[50], which we observe to be significantly reduced in $\text{TUBB}^{\text{G308S}}$ (*lighter green*) compared to $\text{TUBB}^{\text{WT}}$ cells (*darker green*). Importantly, EB1 recruitment has been shown to be affected by alterations in MT structure[51,52].

Migration of the ciliated centrosome to the plasma membrane occurs at a later stage of ciliogenesis and it is driven by pushing forces from MTs nucleation, bundling and polymerization[32]. Consistent with this, the impaired nucleation, altered structure and decreased growth speed of MTs affect this movement in $\text{TUBB}^{\text{G308S}}$ cells.
